# End-to-End Integrative Segmentation and Radiomics Prognostic Models Improve Risk Stratification of High-Grade Serous Ovarian Cancer: A Retrospective Multi-Cohort Study

**DOI:** 10.1101/2023.04.26.23289155

**Authors:** LANCET DIGITAL HEALTH, Kristofer Linton-Reid, Georg Wengert, Haonan Lu, Christina Fotopoulou, Philippa Lee, Federica Petta, Luca Russo, Giacomo Avensani, Murbarik Arshard, Philipp Harter, Mitch Chen, Marc Boubnovski, Sumeet Hindocha, Ben Hunter, Sonia Prader, Joram M. Posma, Andrea Rockall, Eric O. Aboagye

## Abstract

**Background:** Valid stratification factors for patients with epithelial ovarian cancer (EOC) are still lacking and individualisation of care remains an unmet need. Radiomics from routine Contrast Enhanced Computed Tomography (CE-CT) is an emerging, highly promising approach towards more accurate prognostic models for the better preoperative stratification of the subset of patients with high-grade-serous histology (HGSOC). However, requirements of fine manual segmentation limit its use. To enable its broader implementation, we developed an end-to-end model that automates segmentation processes and prognostic evaluation algorithms in HGSOC.

**Methods:** We retrospectively collected and segmented 607 CE-CT scans across Europe and United States. The development cohort comprised of patients from Hammersmith Hospital (HH) (n=211), which was split with a ratio of 7:3 for training and validation. Data from The Cancer Imagine Archive (TCIA) (United States, n=73) and Kliniken Essen-Mitte (KEM) (Germany, n=323) were used as test sets. We developed an automated segmentation model for primary ovarian cancer lesions in CE-CT scans with U-Net based architectures. Radiomics data were computed from the CE-CT scans. For overall survival (OS) prediction, combinations of 13 feature reduction methods and 12 machine learning algorithms were developed on the radiomics data and compared with convolutional neural network models trained on CE-CT scans. In addition, we compared our model with a published radiomics model for HGSOC prognosis, the radiomics prognostic vector. In the HH and TCIA cohorts, additional histological diagnosis, transcriptomics, proteomics, and copy number alterations were collected; and correlations with the best performing OS model were identified. Predicated probabilities of the best performing OS model were dichotomised using k-means clustering to define high and low risk groups.

**Findings:** Using the combination of segmentation and radiomics as an end-to-end framework, the prognostic model improved risk stratification of HGSOC over CA-125, residual disease, FIGO staging and the previously reported radiomics prognostic vector. Calculated from predicted and manual segmentations, our automated segmentation model achieves dice scores of 0.90, 0.88, 0.80 for the HH validation, TCIA test and KEM test sets, respectively. The top performing radiomics model of OS achieved a Concordance index (C-index) of 0.66 ± 0.06 (HH validation) 0.72 ± 0.05 (TCIA), and 0.60 ± 0.01 (KEM). In a multivariable model of this radiomics model with age, residual disease, and stage, the C-index values were 0.71 ± 0.06, 0.73 ± 0.06, 0.73 ± 0.03 for the HH validation, TCIA and KEM datasets, respectively. High risk groups were associated with poor prognosis (OS) the Hazard Ratios (CI) were 4.81 (1.61-14.35), 6.34 (2.08-19.34), and 1.71 (1.10 - 2.65) after adjusting for stage, age, performance status and residual disease. We show that these risk groups are associated with and invasive phenotype involving soluble *N*-ethylmaleimide sensitive fusion protein attachment receptor (SNARE) interactions in vesicular transport and activation of Mitogen-Activated Protein Kinase (MAPK) pathways.

**Funding:** This article represents independent research funded by 1) the Medical Research Council (#2290879), 2) Imperial STRATiGRAD PhD program, 3) CRUK Clinical PhD Grant C309/A31316, 4) the National Institute for Health Research (NIHR) Biomedical Research Centre at Imperial College, London 5) and the National Institute for Health Research (NIHR) Biomedical Research Centre at the Royal Marsden NHS Foundation Trust and The Institute of Cancer Research, London.

**Research In Context:** *Evidence before this study:* Epithelial ovarian cancer (EOC) is the deadliest of all gynaecological cancers, causing 4% of all cancer deaths in women. The most prevalent subtype (70% of EOC patients), high-grade serous ovarian cancer (HGSOC), has the highest mortality rate of all histology subtypes. Radiomics is a non-invasive strategy that has been used to guide cancer management, including diagnosis, prognosis prediction, tumour staging, and treatment response evaluation. To the best of our knowledge, Lu and colleague’s radiomics prognostic vector was the first radiomics model developed and validated to predict overall survival (OS) in HGSOC individuals, from contrast enhanced computed tomography (CE-CT) scans. Both this study and subsequent studies utilised manual segmentations, which adds to the radiologist’s/clinician’s workload and limits widespread use. Additionally, while the models by Lu and co-workers were validated in additional datasets, they were neither harmonised through image resampling – a present requirement for radiomics analysis outlined by the image biomarker standardization initiative – nor compared across machine learning/deep learning models, which could potentially improve predictive performance.

*Added value of this study:* The use of adnexal lesion manually delineated segmentations alone to predict outcome is considered demanding and impractical for routine use. By developing a primary ovarian lesion segmentation, our radiomics-based prognostic model could be integrated into the routine ovarian cancer diagnostic workflow, offering risk-stratification and personalised surveillance at the time of treatment planning. Our study is the first to develop an end-to-end pipeline for primary pre-treatment HGSOC prognosis prediction. Several deep learning and machine learning models were compared for prognosis from CE-CT scan-derived, radiomics and clinical data to improve model performance.

*Implications of all the available evidence:* Our research demonstrates the first end-to-end HGSOC OS prediction pipeline from CE-CT scans, on two external test datasets. As part of this, we display the first primary ovarian cancer segmentation model, as well as the largest comparative radiomics study using machine learning and deep learning approaches for OS predictions in HGSOC. Our study shows that physicians and other clinical practitioners with little experience in image segmentation can obtain quantitative imaging features from CE-CT for risk stratification. Furthermore, using our prognosis model to stratify patients by risk has revealed sub-groups with distinct transcriptomics and proteomics biology. This work lays the foundations for future experimental work and prospective clinical trials for quantitative personalised risk-stratification for therapeutic-intent in HGSOC-patients.

## Introduction

Epithelial ovarian cancer (EOC) is the deadliest of all gynaecological epithelial cancers. High-grade serous histology (HGSOC), is the most prevalent subtype (70% of EOC patients) and confers the highest mortality (1,2). First-line treatment of HGSOC consists of cytoreductive surgery in combination with chemotherapy, such as carboplatin or paclitaxel, with or without bevacizumab (3,4). Several studies have suggested that improvements in response to therapy and prognosis, in clinical trials and current treatment planning, is hindered due to vast degrees of inter and intra-tumoral heterogeneity that traditional molecular markers fail to describe (5).

Radiomics is a non-invasive method that extracts quantitative features from medical imaging in a high-throughput manner (6). Radiomics has been displayed to capture degrees of inter and intra-tumoral heterogeneity and can outperform existing methods in terms of tumour diagnosis and prognosis prediction. For instance, radiomics can predict prognosis in tumours such as head and neck, lung, breast, ovarian cancer, and recently neurodegenerative diseases (7,8).

Previous studies have shown the prognostic potential of radiomics from pre-surgical contrast-enhanced computed tomography (CE-CT) scans in ovarian cancer, and validated in large independent studies (8–10). Importantly, these previous radiomics studies are limited by the requirement to manually segment the lesion of interest. Furthermore, regarding harmonisation and predictive capacity, these prior studies neither involved resampling nor explored multiple machine learning or deep learning techniques. Thus, the earlier radiomics studies warrant broader investigation of various machine learning and deep learning techniques applied in the context of HGSOC automated segmentation and prognosis.

Several feature selection and machine learning techniques exist. However, radiomics studies typically only consider the combination of one feature selection method with one learning algorithm. For instance, van Dijk *et al*., implemented Pearson correlation to identify relevant radiomics features in combination with Lasso regularisation (11). The radiomics study by Lu *et al*., used a univariate Cox regression followed by a Lasso regularised Cox regression (8). It is unclear if these methodological choices yielded models with the highest prognostic accuracy. Thus, a systematic evaluation to identify feature selection methods and learning algorithms is an important step in developing clinically applicable prognostic models.

We propose an end-to-end HGSOC overall survival (OS) prediction pipeline, for which we build several machine learning algorithms that would serve as end-to-end model. These machine learning approaches can either rely on a combination of deep learning for segmentation (Figure 1A) and radiomics (Figure 1B) or a deep learning-based approach without the need for segmentation (Figure 1C). A non-invasive prognostic model based on pre-surgical CE-CT scans could be integrated within the current ovarian cancer diagnostic pathway. This end-to-end tool, in principle, would increase efficiency, reduce manual input, and guide personalised treatment of HGSOC patients.

**Figure 1.**
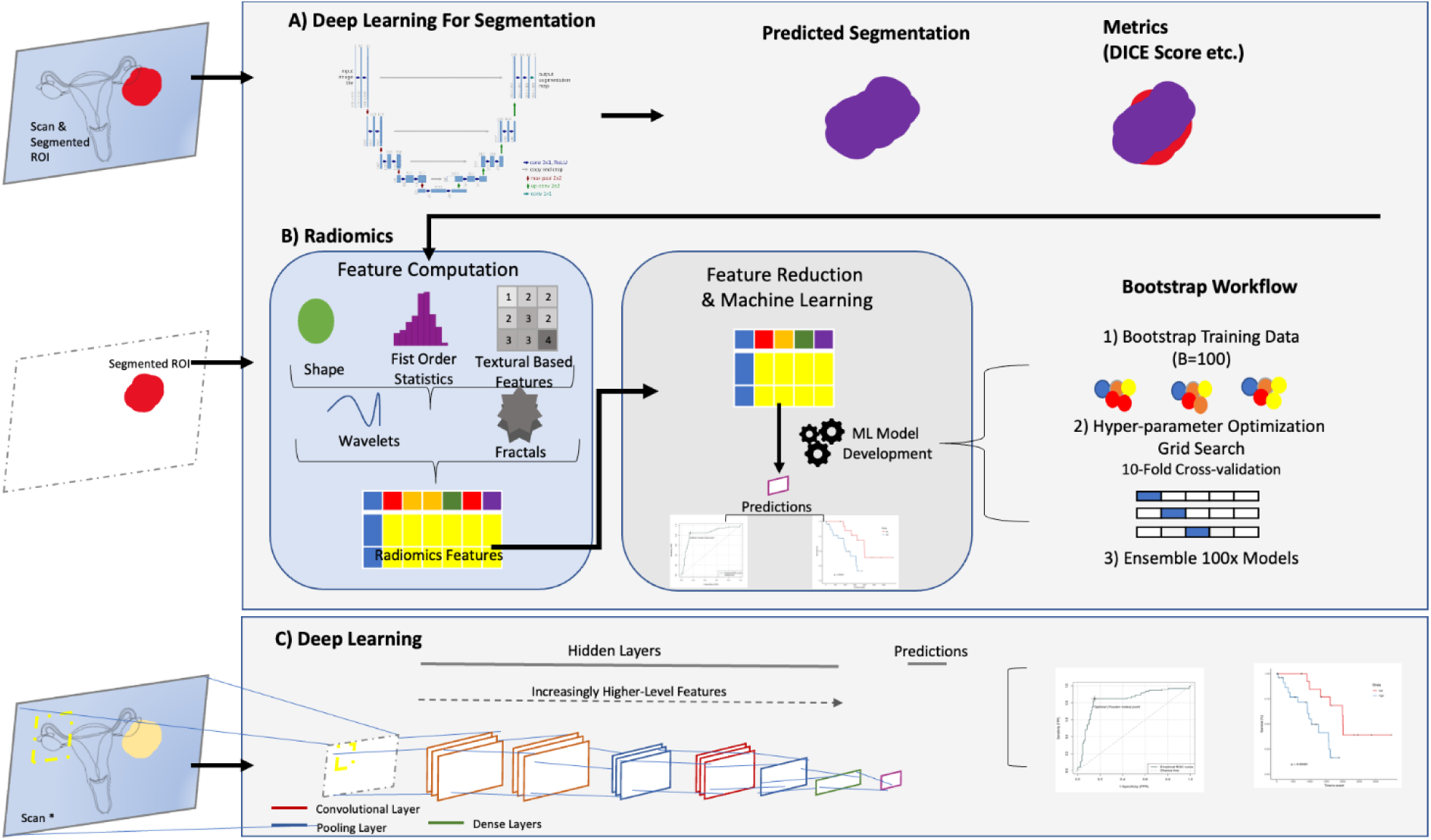
Overview of the AI approach. An end-to-end approach can either rely on a combination of deep learning for segmentation (A) and radiomics (B) or a direct deep learning-based approach without the need for segmentation (C). (A) Illustration of the common approach of using a U-Net-based model to predict segmentation masks together with dice scores as performance metrics. (B) The traditional radiomics feature pipeline from segmented ROI to radiomics feature computation and machine learning modelling. In this work, for each of the feature selection and machine learning algorithm combinations we have bootstrapped to the training dataset 100 times. (C) An alternative to auto-segmentation and radiomics by direct prediction with deep learning – which are often convolutional neural network (CNN) based, for direct end-to-end predictions.

## Methods

### Study design and participating cohorts

In this multi-national retrospective study, we used data from 3 cohorts of primary HGSOC (aged ≥18 years). In the Hammersmith Hospital (HH) cohort (n=211) clinical data (including data related to fresh frozen tissue, imaging, and clinical annotations) were obtained from patients treated at HH between June 2004 and November 2015 (**Table 1**). The HH dataset was split 7:3 into training (n=147), and HH validation (n=64) and used in all machine learning and deep learning models to predict OS, as well as the nn-U-Net segmentation model.

**Table 1.**
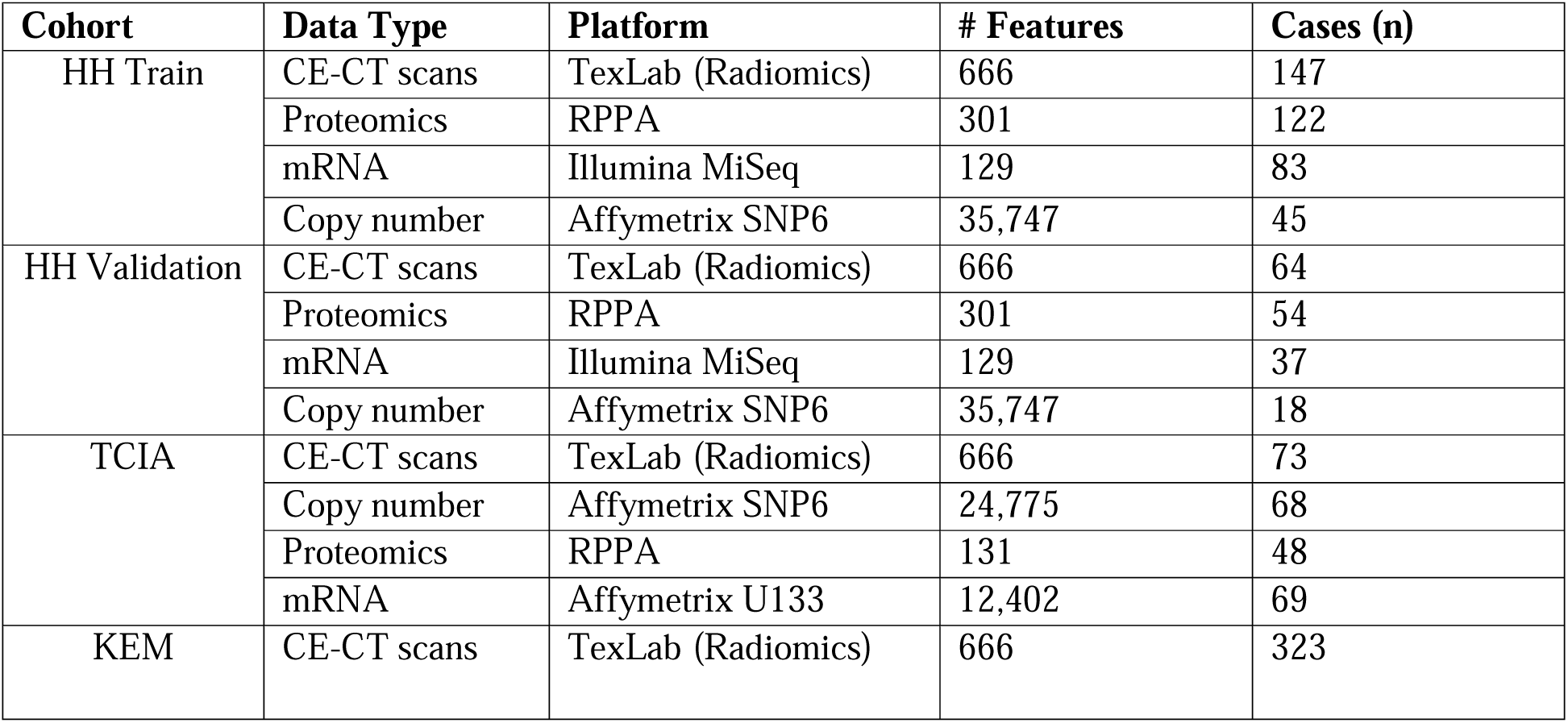
Description of sources of data types used in this study. Hammersmith Hospital (HH) cohort was split 70:30 to HH Train (N=147) and HH validation (N=64). The Cancer Imaging Atlas (TCIA) (N=73) and Kliniken Essen-Mitte (KEM) (N=323) datasets were two hold out test datasets. Proteomics, mRNA, and copy number data were collected in addition to CE-CT scans for the Hammersmith and TCIA datasets. In the KEM dataset only CE-CT scans were collected.

For assessment of our models, we used data from two published benchmark datasets. The first is from The Cancer Imaging Archive (TCIA) cohort (n=73). The second test dataset consisted of 323 cases from Kliniken Essen-Mitte (KEM) and was previously used in (9) to validate the radiomics prognostic vector identified by Lu and colleagues (8).

We used the Radiomics Quality Score (RQS) (12) and Transparent Reporting of a Multi-variable Prediction Model for Individual Prognosis or Diagnosis (TRIPOD) guidelines (https://www.tripod-statement.org/) (Appendix A Tables 1-2) for reporting the development and validation of the prediction models.

### Data Collection and Procedures

A retrospective cohort study was conducted with ethical approval for the analysis of human data, which was obtained from the Hammersmith and Queen Charlotte’s & Chelsea Research Ethics Committee (approval 05/QO406/178) and the Kliniken Essen-Mitte Research Ethics Committee (informed consent was waived). The study was carried out in accordance with the ethical standards of the relevant committees and with the principles of the 1964 Declaration of Helsinki and its subsequent amendments.

The HH training and validation data consisted of patients with HGSOC who received treatment at the Hammersmith Hospital (HH) between June 2004 and November 2015. These patients were selected based on the availability of fresh frozen tumour tissue samples and preoperative CT images. The HH and TCIA cohorts’ patient demographics, surgical and tumour related data were collected retrospectively from medical records and the multidisciplinary team (MDT) notes. Demographic, surgical, and tumour-related data were collected retrospectively from medical records and the multidisciplinary team. With the HH cohort, management of all patients and the indications for surgery were discussed within a multidisciplinary team as per the UK National Health Service (NHS) guidelines. All operations were performed through a midline laparotomy by a specialised dedicated multidisciplinary team within a maximal effort approach aiming to achieve total macroscopic tumour clearance. Standard surgical procedures included peritoneal cytology, extrafascial hysterectomy, bilateral salpingoophorectomy and infra-gastric omentectomy. When indicated, additional procedures, such as dissection of macroscopically suspicious pelvic and paraaortic lymph nodes, bowel resection, splenectomy, diaphragmatic stripping/resection and/or partial resection of other affected organs (e.g., urinary bladder, liver/liver capsule, pancreas, lesser sack) were performed to achieve optimal tumour debulking. No systematic pelvic and paraaortic lymph node dissection was performed routinely in the absence of suspicious bulky lymph nodes (<1 cm). Ninety-seven percent of patients were treated with a platinum-based chemotherapy mainly in a combination regimen with paclitaxel or as monotherapy in isolated cases. Patients were regularly evaluated at the end of their treatment for evidence of disease recurrence. Clinical examination and CA-125 assessment (if the preoperative value was elevated) were performed every 3 months for the first 2 years and then 6-monthly. A CT/MRI-scan was ordered if the above examinations revealed any pathology. An isolated CA-125 increase was not regarded as a recurrence.

In the KEM dataset all consecutive HGSOC patients who underwent primary debulking surgery in between January 2011 and July 2018 (9). Patients were eligible for inclusion if they were confirmed to have HGSOC histology and an evaluable portal venous CT through the primary ovarian tumour mass prior to undergoing upfront cytoreductive surgery, i.e., before any systemic chemotherapy. Any suspicious bulky visible lesions (≥1 cm) from the CT scan were included in the segmentation process. Patients were not eligible if no primary tumour mass was visible due to the initial absence of adnexal mass or due to surgical excision or if the patient has undergone preoperative chemotherapy. All patients in the KEM dataset were to subsequently receive platinum-based combination chemotherapy unless contraindications applied such as poor performance status. Maintenance regimens and availability of clinical trials differed between the KEM and the primary HH study setting, mainly due to funding and licensing differences between the European Medical Agency (EMA) and the UK National Institute for Health and Care Excellence (NICE UK). Oncologic follow-up of the KEM data was performed according to the German follow-up recommendations, mainly symptom-guided and based on clinical and ultrasonographic examination in combination with CA-125 measurement in most patients; initially 3-monthly for the first 3 years and then 6-monthly. A CT or MRI scan was performed if the above examinations revealed any pathology. Isolated CA-125 increase was not regarded as a recurrence. For the HH cohort, follow-up patterns for patient care were similar. Patients were routinely evaluated at the end of their treatment for evidence of disease recurrence. Clinical examination and CA-125 assessment (if the preoperative value was elevated) were performed every 3 months for the first 2 years and then 6-monthly. Even though a CT/MRI scan was ordered if the above examinations revealed any pathology, no routine ultrasonographic examinations were performed at follow-up in asymptomatic patients. Isolated CA-125 increase was not regarded as a recurrence.

The primary outcome measure was OS, defined as the time from the date of surgery until the date of death or last observation. Staging was determined using the FIGO criteria for ovarian epithelial carcinoma (13). In the HH and TCIA dataset optimal debulking was defined as postoperative residual disease less than 10 mm, as this criterion was applied to most of the retrospective patients. Primary chemotherapy resistance was defined as stable disease, partial response, or progressive disease during first-line chemotherapy. Residual disease was dichotomised as either total macroscopic tumour clearance (tumour-free) or the presence of any macroscopic postoperative residual disease (non-tumour-free). A subset of HGSOC patients from TCGA was used as a validation cohort. Preoperative CT images and clinical and histological data for these cases were obtained from the Cancer Imaging Archive and the UCSC Cancer Browser, respectively.

### CE-CT scans, Segmentations and Radiomics Collection

As patients were referred to the cancer centre from a network of cancer units, CE-CT scans were acquired at multiple institutions using different manufacturers and different imaging protocols. The primary tumour masses, of the HH, TCIA, and KEM datasets, were segmented separately by experienced radiologists using ITK snap (Version 3.2, 2015). All segmentations were then checked in consensus with two experienced radiologists specializing in ovarian cancer imaging (GA and AR, with 6- and 16-years’ experience at the time of HH, and TCIA segmentation). The KEM dataset was manually segmented 3 years after HH and TCIA segmentations.

As primary lesions alone harbour prognostic information (8), we did not include secondary lesions in our analysis. As described previously, the entire primary adnexal mass volume (both cystic and solid components) was included in the analysis (8,9). If both adnexa were involved, then both were included in the analysis either as two separate segmentations or as a single segmentation if the mass was confluent. We segmented the entire primary mass including cystic and solid components but excluded ascites. The segmentations only included tissue that was considered highly likely to be cancer by the expert reader. Areas of doubt on CT were not included in any segmentations. In aggregate, inclusion criteria related to the CT images were as follows: primary adnexal mass visible, portal venous phase CT through the adnexal mass, no previous surgical or medical treatment for ovarian cancer. Exclusion criteria related to images were non-contrast or arterial phase CT with no portal venous phase, adnexal mass not included on CT, previous surgery for resection of an adnexal mass, neoadjuvant chemotherapy, the significant artefact for example from metal prostheses that precluded meaningful segmentation of adnexal mass. In accordance with the Image Biomarkers Standardization Initiative (14) CE-CT scans were resampled to isotropic voxel dimensions of 1×1×1 mm.

We developed and compared several different CE-CT, convolutional neural network (CNN) and radiomics based models, independently and in combination with traditional clinical biomarkers, namely age, stage and residual disease. All continuous variables were scaled and mean-centred using training dataset statistics. All categorical variables were ‘one-hot-encoded’.

### Copy Number, Transcriptomics and Proteomics Data Collection

Copy Number, transcriptomics and proteomics data used in this study were previously reported in Lu and colleagues radiomics prognostic vector study (8). For each tumour in the study, one frozen tumour piece was placed into a tube containing 500 μl RLT buffer from RNeasy kit (QIAGEN) and one Retsch 6 mm steel core bead. Tubes were placed into well adapters of a Tissuelyser II (QIAGEN), and tissues were lysed at 15 Hz for 2 min. Tubes were centrifuged briefly and 320 μl was removed for subsequent RNA extraction using the RNeasy kit (QIAGEN) according to the manufacturer’s instructions. RNA concentrations were quantified using the Bioanalyzer system (Agilent).

For DNA extraction, 450 μl of Buffer ATL from the QIAAMP DNA kit (QIAGEN) was added to the centrifuge tube, and DNA was extracted following the manufacturer’s instructions and quantified using QuBit (Thermo Fisher Scientific). Reverse Phase Protein Array (RPPA) arrays were carried out and analysed by MD Anderson Cancer Centre. Here Protein lysates were diluted and loaded onto nitrocellulose-coated slides that had been pre-conjugated with primary antibodies. Each protein was then visualised via a colorimetric reaction and quantified by Array-Pro Analyzer. The raw expression values were then normalised to protein loading and quantified by means of standard curves. Log_2_ transformed and median-centred data were used for the downstream analyses. To perform molecular subtyping, total RNA from each individual case was reverse transcribed into cDNA, followed by amplification with a pool of indexed primers that target a predefined gene list (42 genes). The primers were selected from the Illumina Design Studios. The cleaned PCR product underwent QC by Tapestation (Agilent) to confirm the amplicon sizes. Forty-eight samples were multiplexed in one single MiSeq run. SR 50 bp were used to generate approximately 20 million reads per run.

### Deep learning model development

We developed a U-Net-based segmentation model to facilitate end-to-end application of our models that require segmentation. We base our method on nn-UNet, a recently proposed fully automatic framework for the configuration of U-Net based segmentation methods, that has achieved state-of-the-art performance on 23 public datasets used in international biomedical segmentation competitions (15). We trained the nn-UNet stack of 2D, 3D, and 3D low resolution, and evaluated an ensemble of the four architectures (15).

As there are currently no CNN prognostic models to directly predict OS from CE-CT scans, we built several custom CNNs. ResNet-based architectures were applied without a predefined segmentation region-of-interest (16). ResNet-18, ResNet-32, and a simple CNN were adapted to predict OS by backpropagating the negative loglikelihood loss or the Weibull log loss, similar to that of the Cox proportional hazard and Weibull accelerated failure time model, in the final dense layer. We trained the models for 1,000 epochs with the AdaDelta optimizer and reduce-on-plateau call-back parameters. In the deep learning models to predict OS, we created a cropped around the region of interest, as standard, to prevent exploding or vanishing gradients often caused by non-heterogenous voxel intensity regions.

We developed our radiomics and CNN-based models on radiologist-checked adnexal lesion ground truths as a baseline.

### Radiomics model development

With the 666 textural based radiomics features, computed using TexLab 2.0 (8), we evaluated a combination of 13 different feature reduction techniques with 12 machine learning algorithms, including linear and tree-based techniques, together with boosted and regularised variations. Linearly dependent features were removed, and corresponding lower ordered features were retained.

Several feature selection methods were employed: (I) correlation-based techniques: Pearson, Spearman, Kendall; (II) feature selection algorithms based on mutual information optimisation: joint information maximisation (JIM), mutual information feature selection (MIFS), minimum redundancy maximum relevance (MRMR); and (III) model-based approaches: a univariate Cox-regression model, a random forest variable hunting (RFVH), a random forest variable hunting with maximal depth (RF-MD), a random forest with variable hunting and variable importance (RFVH-VIMP), a random forest with variable hunting and Gini impurity corrected variable importance (IMPRF), and a random forest based on permutation variable importance (PVIRF) (17). Additionally, we also selected features at random for comparative purposes.

For each technique, radiomics feature selection was repeated 100 times by bootstrapping samples of the training cohort. Feature ranks were derived from each of the feature selection technique-specific by averaging the respective technique specific statistics. The number of radiomics features to include was treated as a hyperparameter based on this ranking.

The comparison of different machine learning algorithms included the following non-parametric models: (I) the Cox model, the Cox-Net, and Cox Lasso methods with lasso and elastic-net regularisations; (II) models based on boosting trees (BT): BT_Cox, BT_Cindex; (III) boosting gradient linear models (BGLM): BGLM_Cox, BGLM_Cindex; and (IV) random forest based methods: random survival forest (RSF), random forest using maximally selected rank statistics (MSR_RF), and a random forest with extra trees (ET_RF). Furthermore, we investigated the following full-parametric models (V): survival regression (Survival-Regression) and models based on the Weibull distribution: BT- and BGLM-Weibull.

Hyper-parameter optimisation was performed via grid-search with ten-fold cross-validation, optimising for Concordance index (C-index). Hyper-parameter ranges of the models are listed in Supplementary Table 1. A description of algorithms and feature reduction methods are detailed in the Supplementary Material. A bootstrapping (b=100) strategy was performed at the feature selection and model training stages (Figure 1), and then ensemble models were created. The best model feature importances were computed using the permutation variable importance technique.

### Statistical analysis

To assess the model’s performance for the prediction of OS, we used C-index, hazard ratios (HR), the average time-dependent area-under-the-receiver operating characteristic (tAUC) curve over 1, 2, 3, 4 and 5 years, and Fisher’s exact test P-values. Kaplan-Meier analysis and log-rank tests were applied to assess survival outcomes among sub-groups with pathological complete response or incomplete response.

Quantitative statistics were presented as mean (SD) or median (IQR). Continuous variable distributions between datasets were compared using Kolmogorov-Smirnov (KS) tests, and categorical variables were compared using the χ^2^ test. Spearman Correlation was performed to assess associations between the best model’s predicted probabilities and proteomics, copy number, and transcriptomics data.

The best performing medical image deep learning and radiomics models were selected based on C-index, as it is identical to the area under the curve (AUC) measure. K-means clustering was used on the training dataset derived predicted probabilities from our best performing CE-CT based radiomics model to define risk groups.

### Gene Set Enrichment Analysis (GSEA)

In addition to predicting prognosis in HGSOC patients and performing Kaplan-Meier analysis, we performed GSEA analysis of genes associated with the high-risk group using the clusterProfiler R package. Using univariate logistic regression, we identified genes associated with the high-risk group; in addition we evaluated with the homo sapiens KEGG pathways (hsa), and Genome wide annotation for Human, primarily based on mapping using homo sapiens Entrez Gene identifier-based pathways (ord.HS.ef.db), that are provided by Bioconductor.

## Results

### Clinical Characteristics of data

A total of 618 patients were included in this analysis Patient demographics and clinical parameters are a summarised in Table 2.

**Table 2.** Clinical parameters for combined HH (Hammersmith Hospital) training, HH validation, and The Cancer Imaging Archive (TCIA) and Kliniken Essen-Mitte (KEM) test sets. Categorical data are shown with exact number of subjects (n) per category. Continuous data are summarised with mean and standard deviation (SD) and P-values pertain to Kolmogorov– Smirnov test to compare training and validation cohoorts combines with test cohort distributions. NA = not available.

| Parameter | HH training | HH validation | TCIA | KEM |
| --- | --- | --- | --- | --- |
| Cases (n) | 147 | 64 | 71 | 323 |
| Age: mean (SD) years | 61.85 (11.94) | 61.48 (12.99) | 59.63 (11.10) | 59.74 (11.46) |
| Age: P-value |  |  | 0.14 | 0.036 |
| FIGO stage (n) |  |  |  |  |
| - 1 | 7 | 2 | 6 | 6 |
| - 2 | 9 | 7 | 7 | 14 |
| - 3 | 88 | 39 | 47 | 132 |
| - 4 | 42 | 14 | 11 | 166 |
| - NA | 1 | 2 | 0 | 5 |
| FIGO stage: P-value |  |  | 0.37 | 0.0011 |
| CA-125: mean (SD) U/μl | 1.73 (2.58) | 2.21 (3.11) | NA | 1.11 (1.98) |
| CA-125: P-value |  |  |  | 0.063 |
| Residual disease (n) |  |  |  |  |
| - No | 33 | 18 | 15 | 196 |
| - Yes | 109 | 40 | 50 | 122 |
| - NA | 5 | 6 | 6 | 5 |
| Residual disease: P-value |  |  | 0.99 | 0.99 |
| Primary chemotherapy Response (n) |  |  | NA | NA |
| - Complete Response | 63 | 20 |  |  |
| - Partial response | 19 | 8 |  |  |
| - Progressive Disease | 3 | 3 |  |  |
| - Stable disease | 4 | 7 |  |  |
| - NA | 58 | 13 |  |  |

### nn-UNet Segmentation models

The 2D, 3D-full resolution, and 3D-low resolution nn-UNet model took 81, 130, and 96 hours to train on a 24GB NVIDIA TITAN RTX. We assessed our models using dice score across the HH training, HH validation, TCIA test, and KEM test datasets. Boxplots of the dice scores are shown in Figure 3A. The median scores were 0.96, 0.90, 0.88, and 0.80 for the HH training, HH validation, KEM test, and TCIA test sets, respectively. In Figure 3B we show an example, whereby (in red) the nn-UNet algorithm segmented a bi-lateral lesion, together with the original radiologist segmentation coloured blue. Performance of the 3D-low resolution and 2D are shown in Appendix Figure 1; examples of a high and low performing 3D full-resolution nn-UNet segmentations (dice= 0.89, 0.55) are shown in Appendix Figure 2.

**Figure 3.**
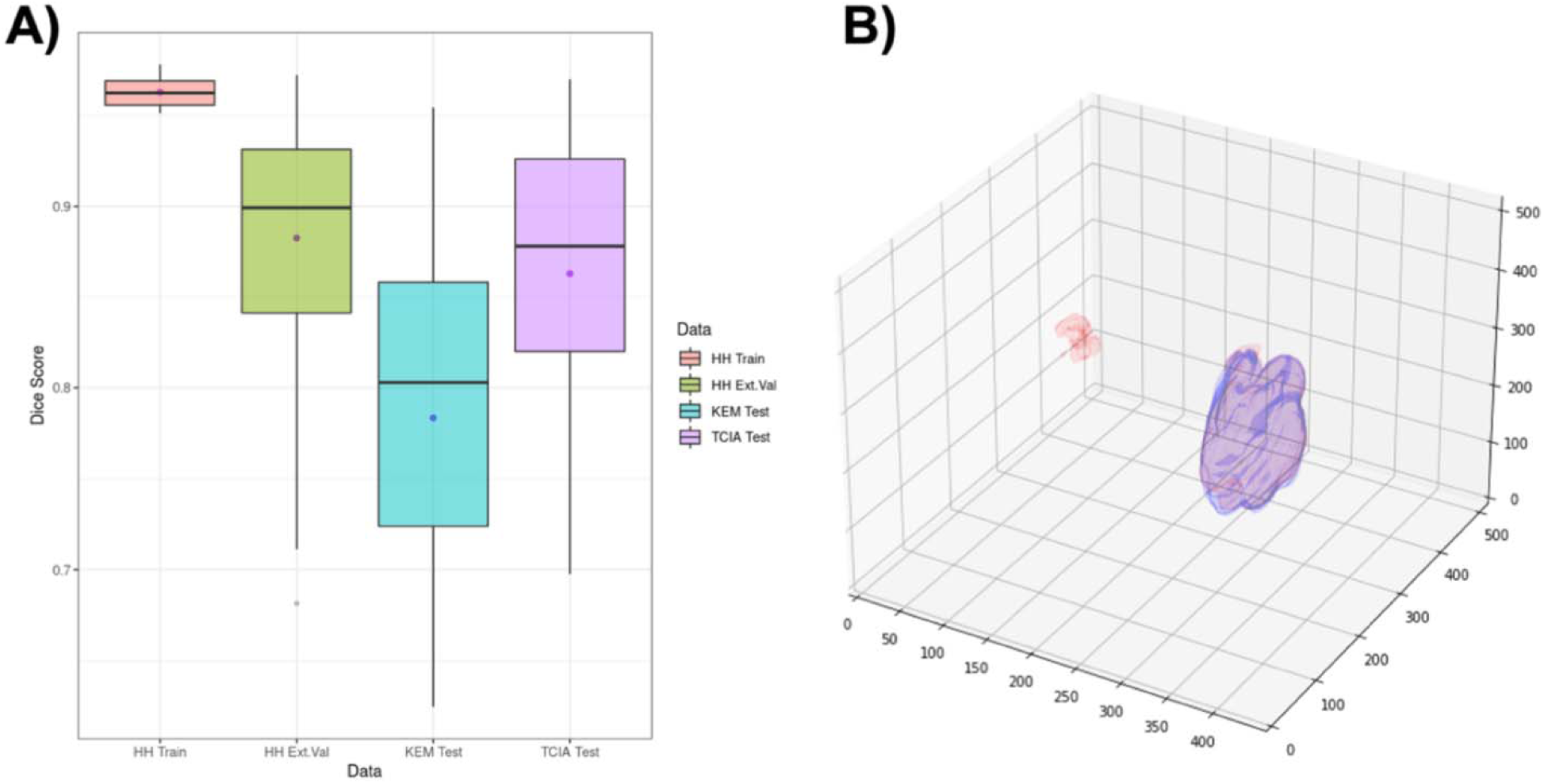
Illustration of 3D high -resolution nn-UNet performance. (A) Boxplots of dice scores for all samples in Hammersmith Hospital (HH) (training (Train) and validation (Val)), Kliniken Essen-Mitte (KEM) and The Cancer Imaging Atlas (TCIA) test datasets. The median scores were 0.96, 0.90, 0.88, and 0.81 for the HH training, HH validation, KEM test, and TCIA test sets, respectively. (B) We show an example: in red is a predicted segmentation and in purple is the original segmentation.

### Preliminary Analysis of Radiomics Data Structure

Data segmented as described above were further used in radiomics analyses. To further understand the radiomic characteristics of the HGSOC subtype, we performed unsupervised Spearman correlation analysis using the radiomic profiles in the HH training cohort (Figure 4A). We show that several radiomics features were highly correlated, and that the CT scanner manufacturer is not a main source of variance (Figure 4B). In the radiomics cohort from HH training cohort we extracted 666 radiomics features from 147 patients performed hierarchal clustering and defined 4 clusters (Figure 4C). Univariate Cox proportional hazard regression shows several radiomics features were significantly associated with OS (Figure 4D), indicating supervised machine learning models would yield value.

**Figure 4.**
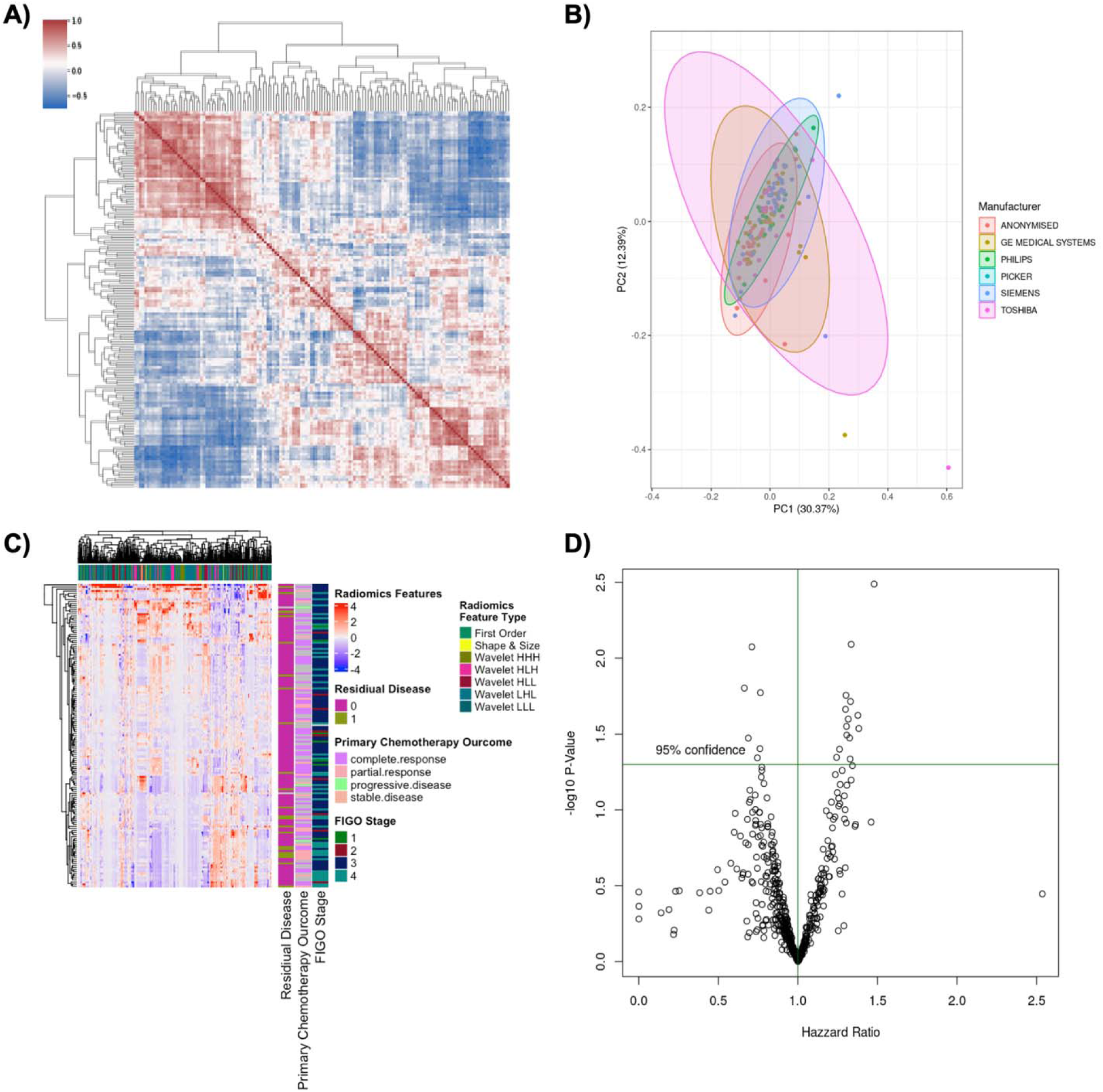
Radiomics Data Structure. (A) Spearman correlation heatmap between radiomics profiles of the training dataset. (B) Principal components 1 and 2 derived from HH training data radiomics highlight the different scanner vendors. (C) Sample-wise radiomics profiles with hierarchal clustering based dendrograms annotated patient-wise (left) and radiomics feature wise (top), radiomics feature type, FIGO stage, CA-125, primary chemotherapy outcome and residual disease annotated respectively. (D) Volcano plot of univariate Cox proportional hazard model log_2_ transformed P-values against the corresponding hazard ratios of the training dataset.

### Supervised Models

In Figures 5A-D, we display the radiomics results for HH training, HH validation, TCIA test, and KEM test sets; for all models used. For OS, the best performing radiomics bootstrap-ensemble model based in terms of external-validation was the Random Survival Forest, built on features selected from the Random Forest-based Feature Selection, using permutation technique described by Janitza and colleagues (18) to calculate the variable importance, i.e. two random forests were implemented, one for feature selection, and a second for prediction on the selected features. We termed this combination – Permutation-Variable Importance Random Forest – Random Survival Forest (PVIRF-RSF). This radiomics model consistently demonstrated high C-index (HH train: 0.85 ± 0.01, HH validation: 0.66 ± 0.06, TCIA: 0.72 72 ± 0.05, KEM: 0.60 ± 0.01). In addition, by dichotomizing the predicted risk (probabilities) from the PVIRF-RSF model using k-means clustering, (threshold, 0.425), we show that this model is capable of stratifying individuals into significantly distinct high and low risk sub-groups, with log-rank P-values: <0.000001, 0.0057, 0.0044, 0.0055, for HH training, HH validation, TCIA and KEM datasets, respectively.

**Figure 5.** Supervised Radiomics Modelling. **(A-D)** Heatmaps of the C-index of different feature selection (x-axis) and modelling strategy (y-axis) for (A) HH training, (B) HH validation, (C) TCIA and (D) KEM. The best performing model (based on HH validation data) was the PVIRF features and RSF model. (E-H) Kaplan Meier curves of the dichotomised predicted probabilities of the PVIRF-RSF (k-means derived threshold of 0.425) of the (E) HH training, (F) HH validation, (G) TCIA and (H) KEM sets.

Whilst k-means clustering showed two distinct high and low risk groups, the number of patients in each group did not capture the most at risk patients. Aiming to derive individuals with highly aggressive phenotypes we also stratified the predictions of PVIRF-RSF model in to three distinct groups termed high, intermediate and low using k-means (k=3 clusters) derived thresholds of 0.33 and 0.44. These risk groups were significantly distinct (P-values: <0.000001, 0.039, 0.014, 0.035) for HH train, HH validation, TCIA, and KEM, respectively) and could capture the 5-30% of patients with increased risk.

When we implement the pipeline (including resampling) for the previous modelling technique that derived the radiomics prognostic vector (Table 5) described in Lu and colleagues (2019) within the cells corresponding to “Cox” on the x-axis and “Cox_Lasso” on the y-axis (Figure 5A-D) we obtain low C-indexes (0.56, 0.52, 0.54, 0.51 for HH train, HH validation, TCIA, and KEM, respectively).

We show univariate Cox Proportional Hazzard performance (C-index, tAUC, and HR) for conventional indicators of HGSOC prognosis, namely age, CA-125, FIGO stage, residual disease *CCNE1*, *BRCA1* and *BRCA2* (copy number variation) for the HH train, HH validation, TCIA and KEM test datasets (Appendix Table 3). The deep learning-based (Simple-CNN, ResNet18, and ResNet34) model performance (C-index, tAUC, and HR) are presented in Appendix Table 4. In Table 4 we compare the PVIRF-RSF model with several traditional clinical models and with the custom-built deep learning-based models (Simple-CNN, ResNet18, and ResNet34).

### Biological interpretation of the radiomics Model

To understand tumour biological characteristics linked to the PVIRF-RSF predictions (Figure 6A), we performed a gene-set enrichment analysis on the corresponding TCIA and HH mRNA data. A volcano plot of the TCIA data with significantly enriched genes highlighted, is displayed in Figure 6B. Here, we found 10 genes to be significantly associated with the high-risk group. We found that Amoebiasis, alpha-Linolenic acid metabolism, and MAPK signalling pathway were the three pathways most significantly enriched for associations with high PVIRF-RF risk groups (P-value= 0.003, 0.003, 0.006). Since several amoebic proteins including lectins, cysteine proteineases, and amoebapores are associated with the invasion process, the results suggest emphasis of an invasive phenotype in the high-risk group. Emphasis of SNARE interactions in vesicular transport, basement membrane, bicellular tight junction and response to fibroblast growth factor (Figure 6 C, D) further support this notion. We show there is a significant increase in Tumour cellularity in the high-risk group the low-risk group (Figure 6 E). In addition, there is a small but insignificant decrease in Rad51 protein expression in the high-risk group compared to the low-risk group within the HH cohort (Figure 6 F), although there is a small significant decrease in Rad51 protein expression in the high-risk group compared to the low-risk group in the TCIA data set (Appendix Figure In the TCIA proteomics data, the features that were negatively correlated JNK2, YB1, RAD51, MTORPS448, and HER3PY1298 were the most correlated with the predictions. The most significantly associated protein, STAT5ALPHA was positively correlated with the predicted probabilities (Figure 7 and supplementary Table 3).

**Figure 6.**
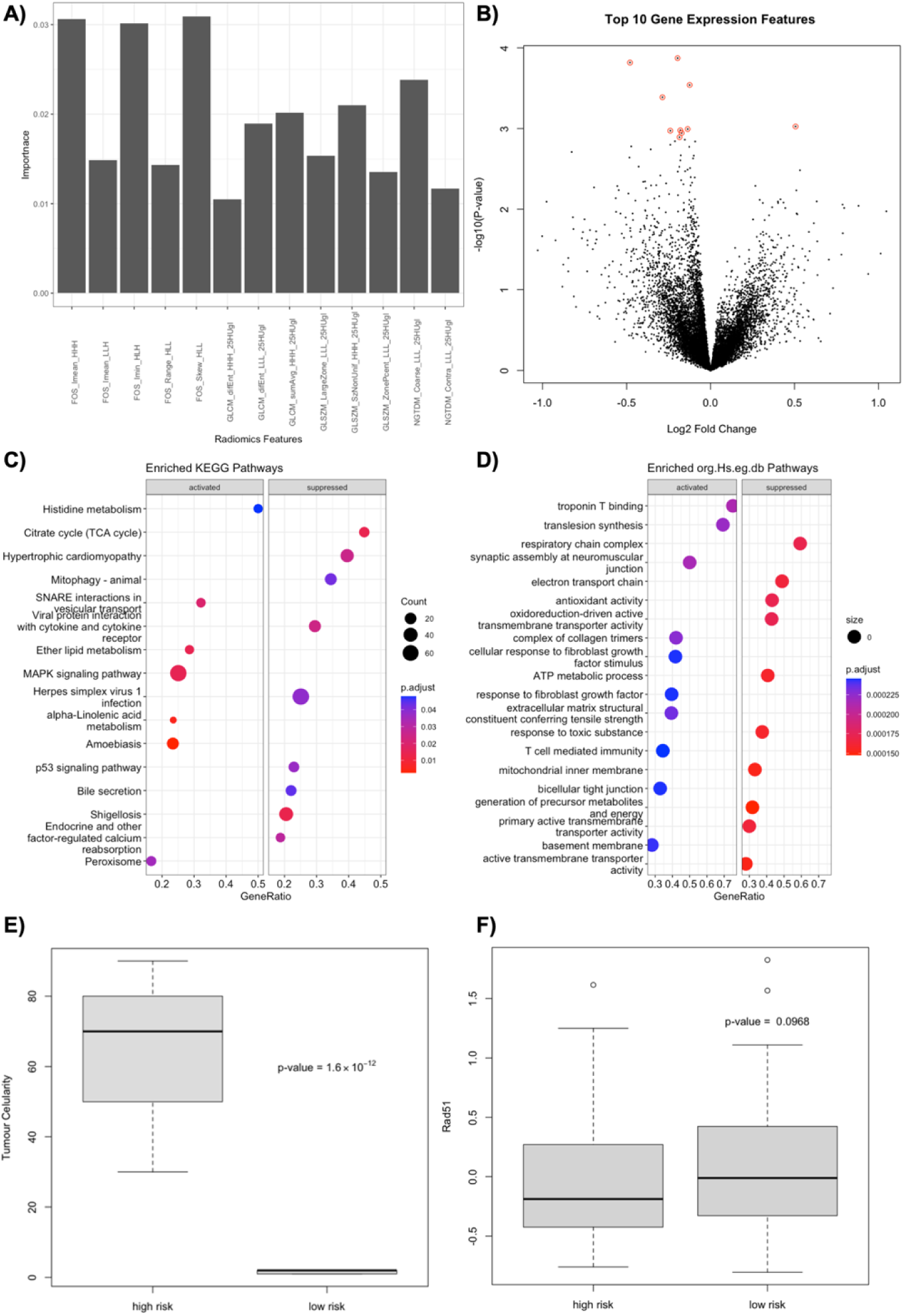
Volcano Plot and GSEA of Permutation-Variable Importance Random Forest – Random Survival Forest (PVIRF-RSF) Model. (A) Radiomics features and there premutation computed importance in the PVIRF-RSF model (B) Volcano plot of genes expressed that are associated with the predicted probabilities of PVIRF-RSF radiomics model (TCIA data). (C) Top 10 KEGG based activated and suppressed pathways probabilities of PVIRF-RSF (TCIA data). (D) Top 10 Genome wide annotation for Human, primarily based on mapping using Entrez Gene identifiers (ord.HS.ef.db) activated and suppressed pathways probabilities of PVIRF-RSF (TCIA data). (E) HH data Tumour cellularity box plot across risk groups. (F) HH data Rad51protein expression across PVI-RSF risk groups.

**Figure 7.**
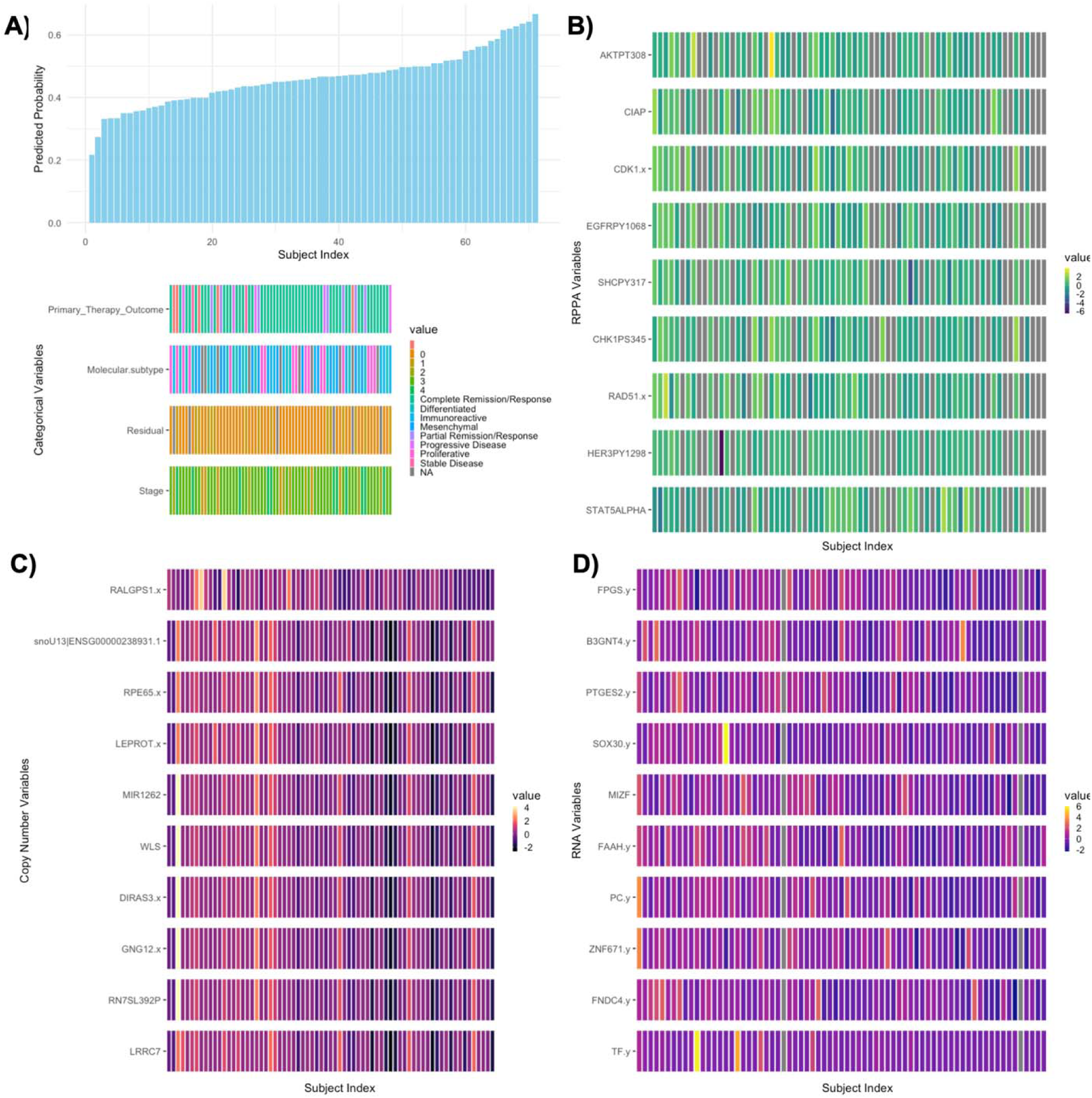
Univariate Analysis Testing for key variables associated with predictions of the Permutation-Variable Importance Random Forest – Random Survival Forest (PVIRF-RSF) Model Derived from TCGA data. (A) The predicted probabilities for all 71 subjects (x-axis) in the TCIA cohort based on inference from the PVIRF-RSF Model and Clinical Variables. (B) Top ten RPPA variables most correlated with predictions of PVIRF-RSF (x-axis corresponds to each sample). (C) Top ten RNA variables most correlated with predictions of PVIRF-RSF (x-axis corresponds to each sample). (D) Top ten Copy Number variables most correlated with predictions of PVIRF-RSF (x-axis corresponds to each sample).

In the TCIA copy number variation data, the genes *LRRC7, RN7SL392P, GNG12.x, DIRAS3, WLS, MIR1262, RPE65, LEPROT, snoU13|ENSG00000238931.1, RALGPS1* were the most correlated (negative correlation) (Figure 7 and Appendix Table 4). In the TCIA RNA expression data, the genes *TF, FNDC4, ZNF671, PC, FAAH, MIZF, SOX30, PTGES2, B3GNT4, FPGS* were the most correlated (negative correlation) (Figure 7 and Appendix Table 4).

In https://data.mendeley.com/xxx we provide the corresponding coefficients, Spearman Rho coefficients, and adjusted P-values. In the HH cohort we display a significant increase in tumour cellularity in the high-risk group compared to the low-risk group (Supplementary Figure 4).

The study by Lu *et al*. (2019) suggested that RPV emphasised an activated stromal phenotype. From the foregoing, demonstrate that the new model, PVIRF-RSF, emphasised different biology, we compared examined its relationship to histology data. PVIRF-RS is strongly positively correlated with tumour cellularity and not fibronectin indicating marked differences between the two models, with the PVIRF-RS emphasising the tumour (invasive) rather than the stromal compartment.

## Discussion

The mesoscopic architecture of ovarian cancer contains mineable features that can provide prognostic and predictive tools for patient management. We present here an advanced analytical approach for mining such data, together with new biological and biophysical insights not previously appreciated, and clinical utility of the approach.

Almost all patients with ovarian cancer will undergo imaging including CE-CT prior to surgical or non-surgical treatment. In previous work by Lu *et al*. (2019), a radiomics model that is strongly related to tumour stroma was disclosed (8). This model, RPV, was found to be robust in a subsequent large European validation study (9). This earlier work supports the development of novel technical approaches to harness the vast information and to enable an appreciation of new biology at a scale beyond that of microscopy – the mesoscopic scale – with radiomics alone (8) or combined with histopathology, genetic and clinical factors (19). Assessing primary ovarian cancer CE-CT is an iterative and labour-intensive process, even to expert radiologists but not computers. In this regard, radiomics and CNN algorithms, once developed, can process massive quantities of image data efficiently, are not vulnerable to fatigue, and have high throughput and stability. We present, for the first time, an end-to-end tool that automatically segments ovarian primary mass and quantifies radiomics signatures to predict prognosis.

Distinct from previous radiomics work (8), we offer a tool to segment the primary ovarian mass and produce prognostic and biological information in a full end-to-end machine learning pipeline. We built and validated 156 radiomics based models across 100 bootstrapped training datasets in a grid-search cross-validation setting to predict OS using time-to-event outcomes. We compared these radiomics models with a) deep learning CNN based models, and b) clinical baselines, namely CA-125 and transcriptomics subtypes (20,21) to assert superiority and ensure that the tool model will be widely used when reported. By means of evaluating the best C-index, PVIRF-RSF was identified as the best performing modelling when applied to validation data, exceeded performance of clinical data and deep learning architectures (Figure 5, Table 4, Table 5). While several studies have displayed success of deep learning for classification with CNNs, there remains to date relatively little literature on using CNNs for survival analysis. The performance of the deep learning-based CNN models is lower than the top performing radiomics model in the present study. Of note, however, we only investigated three different architectures; recently several feed-forward networks have been proposed to predict survival (22–25). Future work on adapting CNNs with various functions to model survival will help identify whether there could be unseen benefits from CNNs in this setting.

**Table 3.**
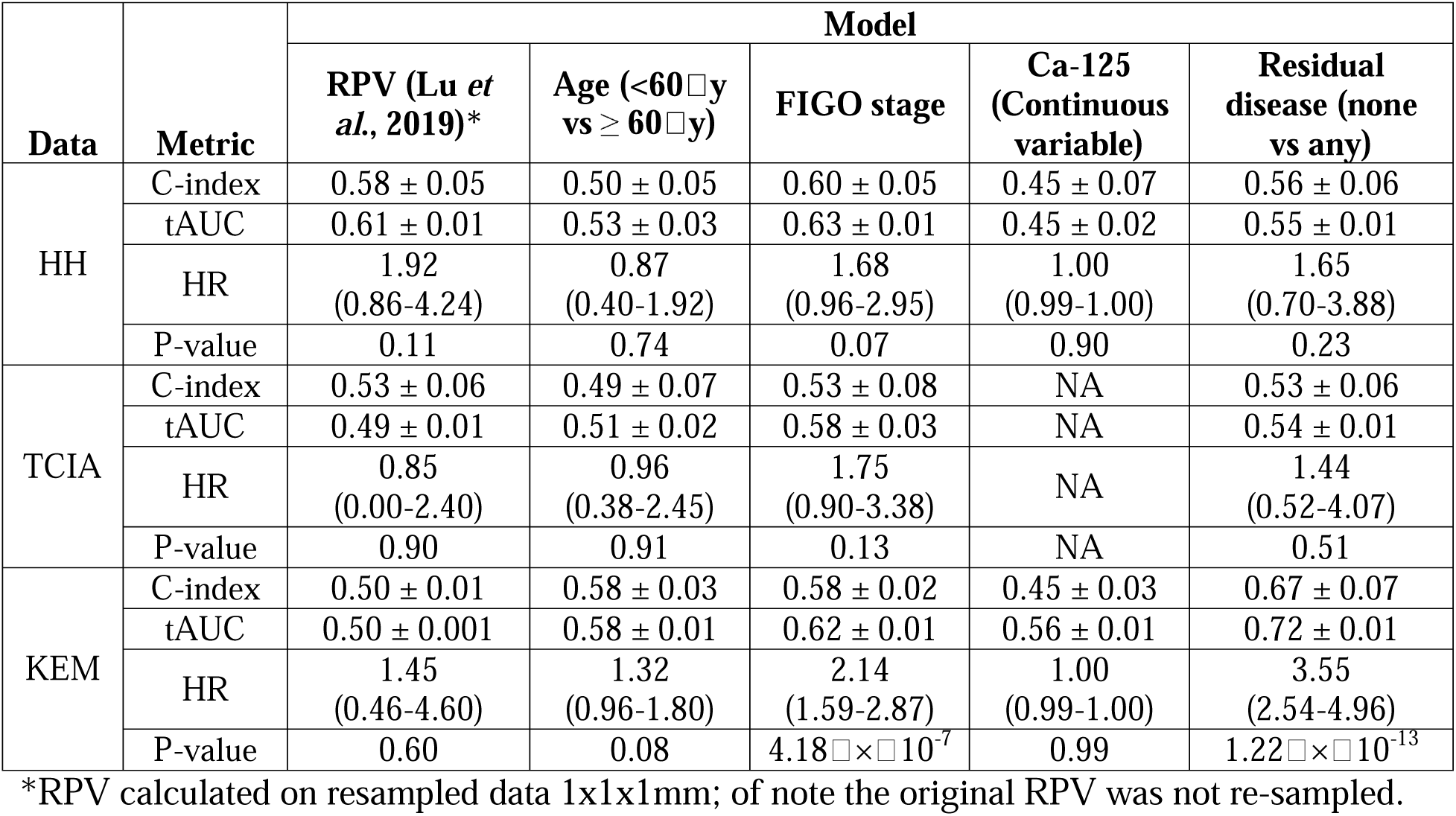
Performance of traditional radiomics (RPV) and clinical baseline univariate Cox proportional hazards (CoxPH) models. . A summary of performance using C-index and standard error, average time-dependent area-under-the-receiver operating characteristic curve (tAUC) over 1, 2, 3, 4 and 5 years with standard error, Hazard ratio (HR), and with 95% confidence intervals. Performance metrics for RPV, age, FIGO stage, Ca-125, and residual disease. NA = not available.

| Data | Metric | Model |  |  |  |  |
| --- | --- | --- | --- | --- | --- | --- |
|  |  | RPV (Lu *et al.*, 2019)* | Age (<60 □ y vs ≥ 60 □ y) | FIGO stage | Ca-125 (Continuous variable) | Residual disease (none vs any) |
| HH | C-index | 0.58 ± 0.05 | 0.50 ± 0.05 | 0.60 ± 0.05 | 0.45 ± 0.07 | 0.56 ± 0.06 |
|  | tAUC | 0.61 ± 0.01 | 0.53 ± 0.03 | 0.63 ± 0.01 | 0.45 ± 0.02 | 0.55 ± 0.01 |
|  | HR | 1.92<br>(0.86-4.24) | 0.87<br>(0.40-1.92) | 1.68<br>(0.96-2.95) | 1.00<br>(0.99-1.00) | 1.65<br>(0.70-3.88) |
|  | P-value | 0.11 | 0.74 | 0.07 | 0.90 | 0.23 |
| TCIA | C-index | 0.53 ± 0.06 | 0.49 ± 0.07 | 0.53 ± 0.08 | NA | 0.53 ± 0.06 |
|  | tAUC | 0.49 ± 0.01 | 0.51 ± 0.02 | 0.58 ± 0.03 | NA | 0.54 ± 0.01 |
|  | HR | 0.85<br>(0.00-2.40) | 0.96<br>(0.38-2.45) | 1.75<br>(0.90-3.38) | NA | 1.44<br>(0.52-4.07) |
|  | P-value | 0.90 | 0.91 | 0.13 | NA | 0.51 |
| KEM | C-index | 0.50 ± 0.01 | 0.58 ± 0.03 | 0.58 ± 0.02 | 0.45 ± 0.03 | 0.67 ± 0.07 |
|  | tAUC | 0.50 ± 0.001 | 0.58 ± 0.01 | 0.62 ± 0.01 | 0.56 ± 0.01 | 0.72 ± 0.01 |
|  | HR | 1.45<br>(0.46-4.60) | 1.32<br>(0.96-1.80) | 2.14<br>(1.59-2.87) | 1.00<br>(0.99-1.00) | 3.55<br>(2.54-4.96) |
|  | P-value | 0.60 | 0.08 | 4.18 □ × □ 10<sup>-7</sup> | 0.99 | 1.22 □ × □ 10<sup>-13</sup> |

\*RPV calculated on resampled data 1x1x1mm; of note the original RPV was not re-sampled.

**Table 4.** Performance of Radiomics PVIRF-RSF, and deep learning-based models.

| Data | Metric | Model |  |  |  |  |
| --- | --- | --- | --- | --- | --- | --- |
|  |  | PVIRF-RSF<br>(continuous<br>variable) | Simple CNN | ResNet-18 | ResNet-34 | PVIRF-RSF<br>(categorical<br>variable) |
| HH | C-index | 0.66 ± 0.042 | 0.58 ± 0.05 | 0.56 ± 0.06 | 0.52 ± 0.04 | 0.64 ± 0.05 |
|  | tAUC | 0.73 ± 0.011 | 0.40 ± 0.01 | 0.55 ± 0.02 | 0.52 ± 0.02 | 0.64 ± 0.01 |
|  | HR | 0.0081<br>(0.00010-0.38) | 0.99 (0.99-1) | 0.99 (0.9-1) | 0.99(0.99 -<br>1.1) | 3.84 (1.43-10.3) |
|  | P-value | 0.010 | 0.62 | 0.51 | 0.52 | 0.008 |
| TCIA | C-index | 0.72 ± 0.05 | 0.53 ± 0.081 | 0.56 ± 0.07 | 0.55 ± 0.05 | 0.73 ± 0.05 |
|  | tAUC | 0.78 ± 0.010 | 0.49 ± 0.01 | 0.59 ± 0.02 | 0.53 ± 0.02 | 0.74 ± 0.01 |
|  | HR | 0.00062<br>(0.0000013-0.3) | 0.26<br>(0.0025- 1) | 1.05<br>(0.00011-1.1) | 0.99(0.00011<br>-1.1) | 3.69 (1.47-9.6) |
|  | P-value | 0.021 | 0.62 | 1.00 | 1.00 | 0.0074 |
| KEM | C-index | 0.60 ± 0.03 | 0.52 ± 0.05 | 0.55 ± 0.05 | 0.52 ± 0.04 | 0.57 ± 0.02 |
|  | tAUC | 0.60 ± 0.0001 | 0.49 ± 0.01 | 0.55 ± 0.02 | 0.55 ± 0.02 | 0.56 ± 0.01 |
|  | HR | 0.038 (0.0041-0.40) | 0.26<br>(0.001- 1) | 0.42<br>(0.031- 1.10) | 0.99<br>(0.99- 1.10) | 1.83 (1.18 -<br>2.82) |
|  | P-value | 0.0070 | 0.84 | 0.91 | 0.91 | 0.0065 |

**Table 5.** Performance of Radiomics Multi-variable PVIRF-RSF (continuous variable) and additional clinical variables)

| Data |  | HH | TCIA | KEM |
| --- | --- | --- | --- | --- |
| C-index |  | 0.73 ± 0.07 | 0.73 ± 0.06 | 0.73 ± 0.03 |
| tAUC |  | 0.76 ± 0.02 | 0.78 ± 0.03 | 0.75 ± 0.00 |
| P-value |  | 0.02 | 0.043 | 1.01 × 10<sup>-12</sup> |
| HR | Age | 0.99 (0.96-1.02) | 1.04 (0.98-1.10) | 1.02 (0.99-1.03) |
|  | Stage | 2.08 (1.08 -4.05) | 1.70 (0.78-3.71) | 2.10 (1.38-3.21) |
|  | Residual disease | 1.26 (0.52-2.03) | 1.06 (0.30-3.70) | 3.61 (2.23-5.64) |
|  | PVIRF-RSF | 0.0042 (7.1×□<br>10-5 -0.24) | 0.00016 (2.17×□<br>10-7 - 0.12) | 0.04 (0.00-0.05) |

**Table 6.**
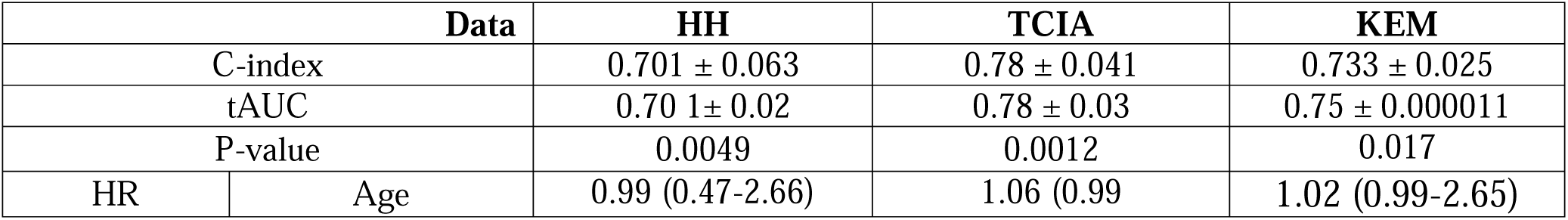
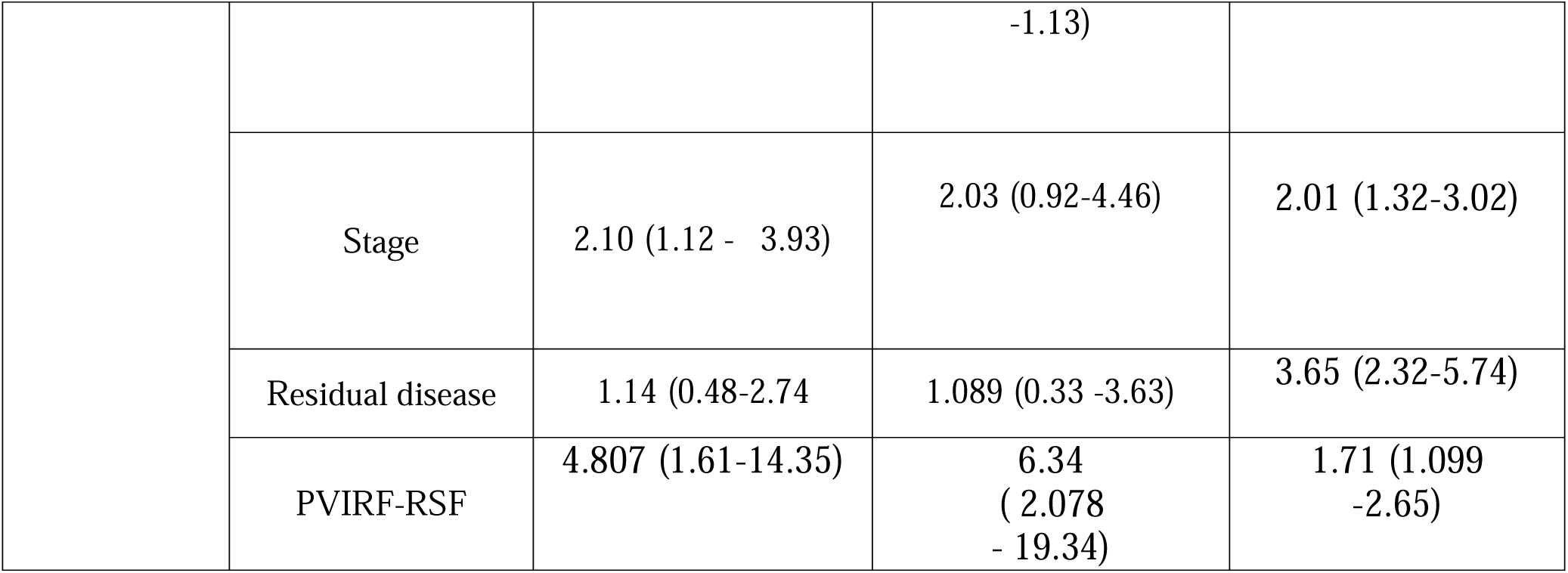
Performance of Radiomics Multi-variable PVIRF-RSF (categorical variable) and additional clinical variables)

Biophysically, the PVIRF-RSF vector comprised of 13 radiomics features (Figure 6A) including discrete wavelet transformation-derived features. Our model directly shared only one feature with the RPV described by Lu and colleagues (FOS_Imedian_LHH). Otherwise, the majority of radiomics features used in this study are different from that of the RPV model (8). In particular the features suggest emphasis of skewed structures and contrast (e.g. FOS_Skew_HLL depicting asymmetry of distribution around the mean, and NGTDM_Contra_LLL_25HUgl depicting contrast - the difference between a grey value and the average grey value of its neighbours within a distance). This led us to investigate the biology emphasised by the PVIRF-RSF model. An interesting finding is that genes and pathways involved in a tumour’s invasive potential were emphasised. Regarding genes and pathways, SNAREs, emphasised in high PVIRF-RSF stratified groups, facilitate intracellular vesicle trafficking, including that involved in the transport of invadopodium-associated proteins, and in so doing promote modification of ECM and modulation of signalling pathways involved in the movement of cancer cells (26). Genes involved in response to fibroblast growth factor stimulus, extracellular matrix constituents, basement membranes and bicellular tight junctions were associated with high PVIRF-RSF. In keeping with above the high-risk PVIRF-RSF stratified group was associated with high cellularity, high expression of the MAPK signalling. In this setting, expression of RAD51, a marker of DNA repair capacity, was found to be slightly lower (Figure 6, Supplementary) in the high-risk group, distinct from the reported poor prognosis of RAD51 (29) The exact biology emphasised by PVIRF-RSF remains incomplete and future efforts will aim to elaborate this.

Regardless of the exact biology we show that PVIRF-RSF has higher C-index than any other vector for ovarian cancer. The categorisation of the PVIRF-RSF predictions through k-means facilitates stratification (threshold 0.425) of HGSOC patients into “high” and risk “low” groups with high HR (Table 4). The median overall survival of individuals in the high and low-risk group were 13, 17, 10, and 14 months and 30, 29, 21, 22 months for the HH training, HH validation, TCIA and KEM datasets, respectively, thus 2-3 times shorter. After adjusting the PVIRF-RSF defined high-risk group for age, stage, status and residual disease, the Hazard ratios were 4.81 (1.61-14.35), 6.34 (2.08-19.34), and 1.71 (1.10-2.65). This supports the potential clinical use of a model that can be embedded in CE-CT workstations to provide radiologists and gynaecological oncologists with real-time, segmentation, prognosis, and tumour environment information. Furthermore, the use of two test datasets suggests the model generalizes well, and can be used internationally, subject to further validation, to help alleviate the uneven distribution of medical and human resources. The segments may also be used to provide other imaging biomarker information, for example when combined with RPV (8). The PVIRF-RSF high group represent patients likely to benefit less from standard of care surgery combined with chemotherapy. Whether these patients will benefit from upfront anti-invasive therapy together with surgery will be a future hypothesis to be tested.

Previous studies have defined molecular features associated with prognosis from biopsy-based data such as gene expression, DNA methylation, CNA, and more recently microRNA and circulating tumour DNA(26,27,29). However, biopsy-derived data and subsequent models are often challenging to translate into routine clinical use due to the invasiveness of a biopsy, insufficient prognostic power due to the vast intratumor heterogeneity, high assay costs, and reproducibility constraints. Currently, treatment may be performed without any understanding of an individual’s prognosis; our approach would facilitate early estimation of these characteristics. The prognostic model we have developed is derived from a patient’s preoperative CE-CT scan at the presentation of the disease and could be integrated with the current clinical pipeline. The integrative segmentation and PVIRF-RSF radiomics model based on CE-CT enables a non-invasive interpretable state-of-the-art prognostic evaluation of HGSOC. Our predictive model, in principle, improves the risk stratification of patients with HGSOC. Our methods represent a unique future framework for producing more granular, individualised, and robust prognostic models. We have described a crucial technology that could breach the unmet need to rapidly define prognosis in HGSOC patients and facilitate rapid patient entry into clinical trials at the point of care.

## Data Availability

All data produced in the present study are available upon reasonable request to the authors

## Contributors

EOA, JMP and KL-R, AR and CF designed the study. KL-R, AR, GW, CF, and HL were responsible for data collection. AR, GA, GW, and MA completed the radiologist segmentation on the HH and TCIA cohorts. AR, CF, GA, GW, HL assessed and characterised the HH and TCIA cohorts. HL collected the proteomics, transcriptomics and copy number data. KLR and HL extracted the radiomics and copy number, transcriptomics count data and proteomics features. KLR, EOA, JMP, AR contributed to discussions of this study at weekly lab meetings. CF, SP, PH, PL, LR, and AR was responsible for providing the KEM dataset and radiological segmentations. KL-R, MB, SH, BH, MC and contributed to statistics and machine learning. KL-R, EOA and JMP wrote the manuscript. All authors reviewed and approved the submitted manuscript.

## Appendix

**Figure 1.**
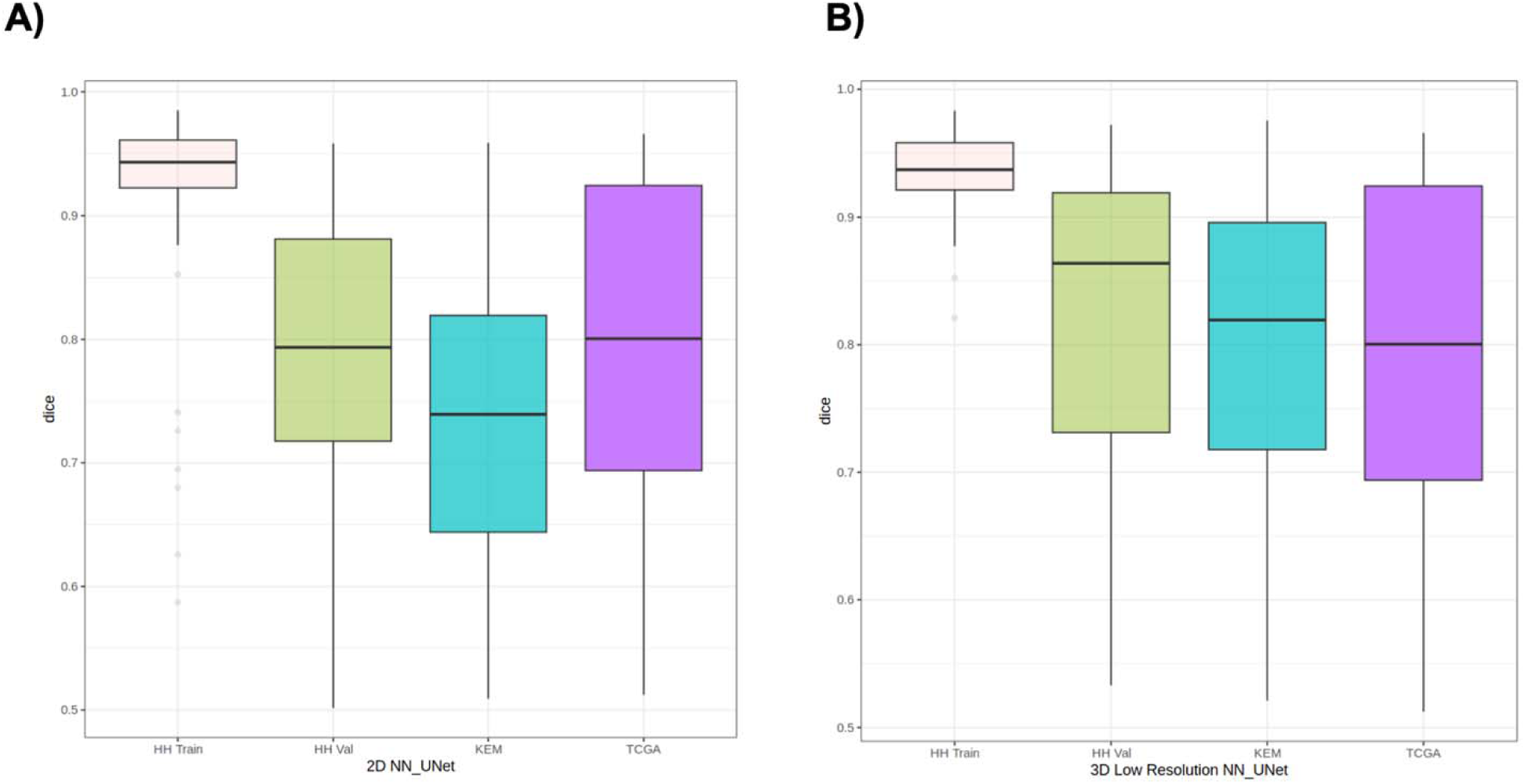
Illustration of 2D and 3D low -resolution nn-UNet performance. Boxplots of dice scores for all samples in Hammersmith Hospital (HH) (training (Train) and HH validation (HH.Val)), Kliniken Essen-Mitte (KEM) and The Cancer Imaging Atlas (TCIA) test datasets for (A) 2D nn-UNet and (B) 3D low-resolution nn-UNet.

**Figure 2.**
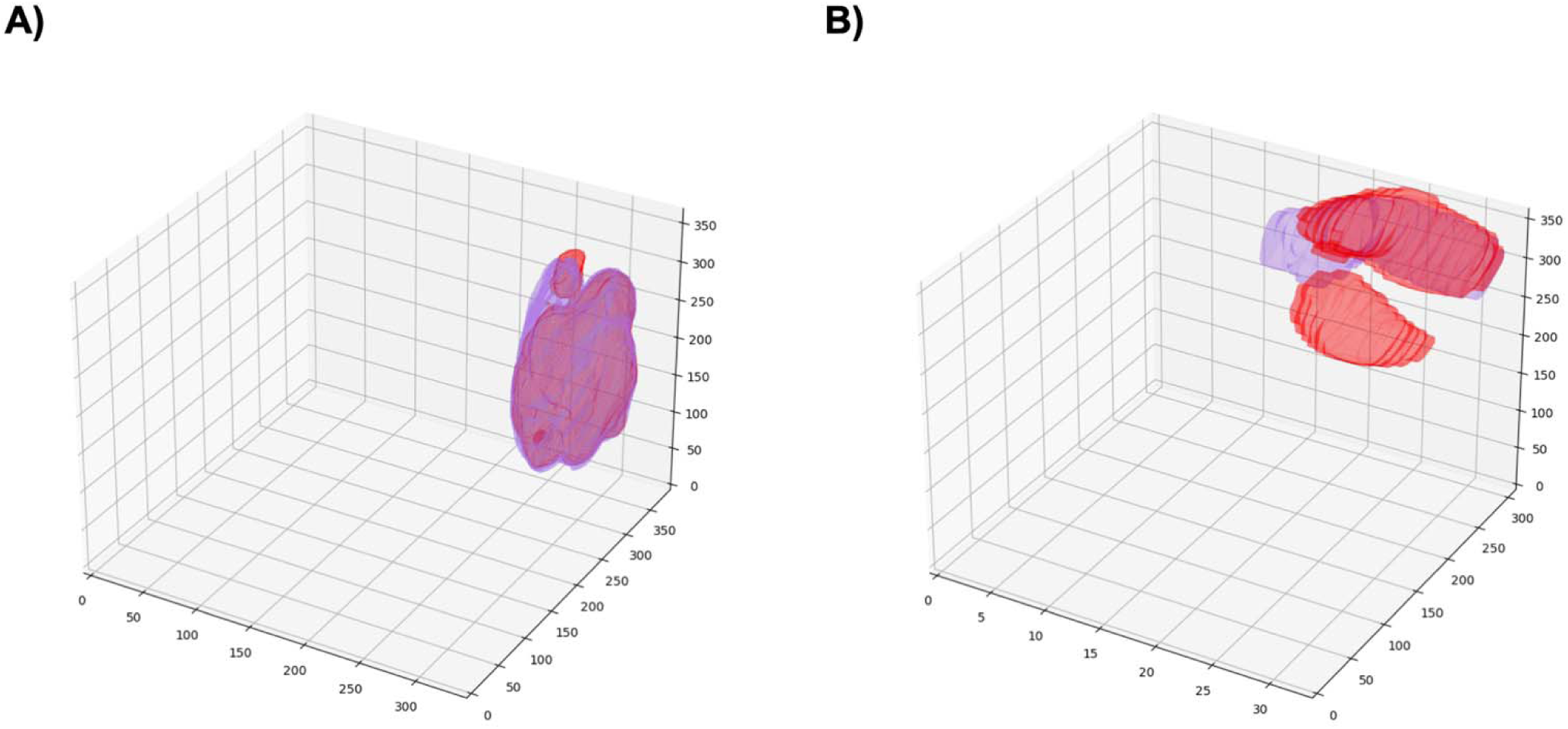
Illustration of 2D and 3D high-resolution nn-UNet segmentation. (A) Mesh plot of the nn-UNet segmentation (red) and manual segmentation (purple) with a Dice score of 0.89. (B) Mesh plot of a nn-UNet segmentation (red) and manual segmentation (purple) with a dice score of 0.55.

**Figure 3.**
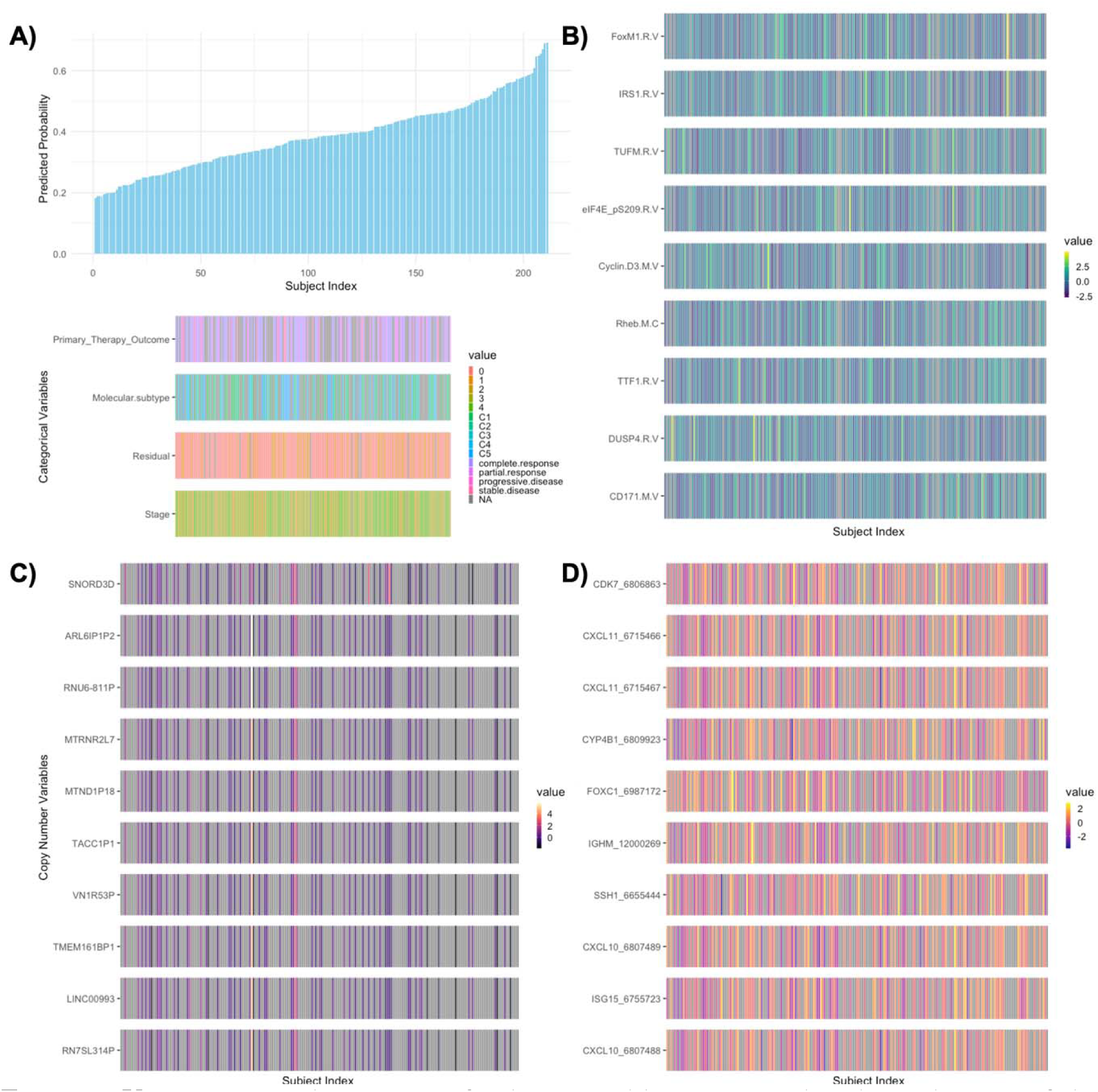
Univariate analysis testing for key variables associated with predictions of the Permutation-Variable Importance Random Forest – Random Survival Forest (PVIRF-RSF) model derived from HH data. (A) The predicted probabilities for all 71 subjects in the Cohort based on inference from the PVIRF-RSF Model and Clinical Variables, ordered by probability. (B) Top ten RPPA variables most correlated with predictions of PVIRF-RSF (ordered in the same manner). (C) Top ten RNA variables most correlated with predictions of PVIRF-RSF. (D) Top ten copy number variables most correlated with predictions of PVIRF-RSF. Colour maps correspond to normalised values of the respective data sets.

**Figure 4.**
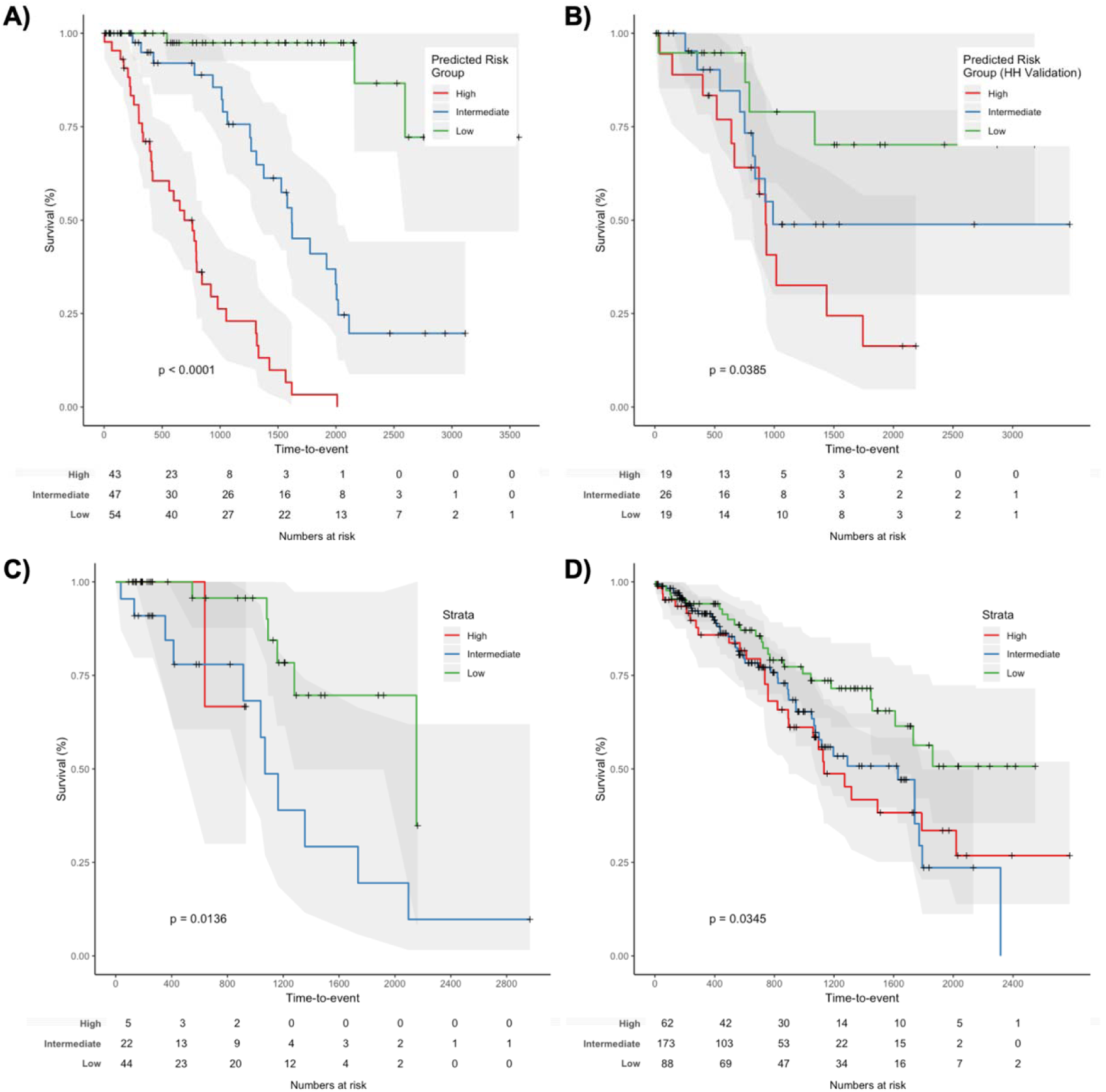
In (A-D) we show Kaplan-Meier curves of the dichotomised predicted probabilities of the PVIRF-RSF (k-means derived thresholds of (0.334), (0.442) for the HH training, HH validation, TCIA and KEM test sets, respectively.

**Figure 5.**
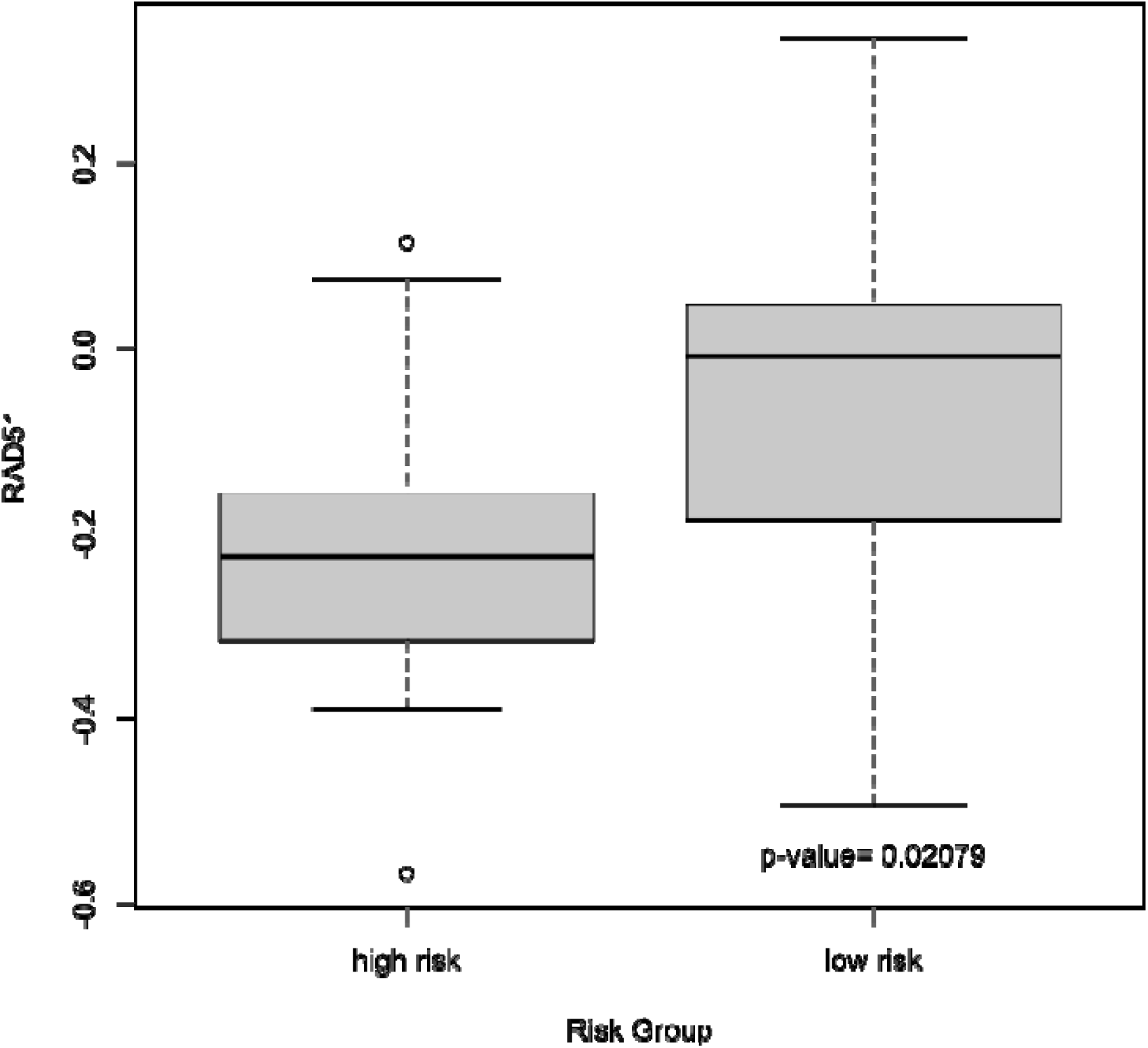
RAD51 protein expression in the TCIA Cohort. Compared across the two risk groups defined with threshold of 0.425.

**Table 1.** **The Radiomics Quality Score** for our study is tabulated below. Our study scores 18/36 on the RQS.

| Criteria | Points | Our Score |
| --- | --- | --- |
| 1. Image protocol quality - well-documented image protocols (for example, contrast, slice thickness, energy, etc.) and/or usage of public image protocols allow reproducibility/replicability | + 1 (if protocols are well-documented) + 1 (if public protocol is used) | 0 |
| 2. Multiple segmentations - possible actions are: segmentation by different physicians/algorithms/software, perturbing segmentations by (random) noise, segmentation at | + 1 | 0 |
| different breathing cycles. Analyse feature robustness to segmentation variabilities |  |  |
| 3. Phantom study on all scanners - detect inter-scanner differences and vendor-dependent features. Analyse feature robustness to these sources of variability | + 1 | 0 |
| 4. Imaging at multiple time points - collect images of individuals at additional time points. Analyse feature robustness to temporal variabilities (for example, organ movement, organ expansion/shrinkage) | + 1 | 0 |
| 5. Feature reduction or adjustment for multiple testing - decreases the risk of overfitting. Overfitting is inevitable if the number of features exceeds the number of samples. Consider feature robustness when selecting features | – 3 (if neither measure is implemented) + 3 (if either measure is implemented) | 3 |
| 6. Multivariable analysis with non-radiomics features (for example, EGFR mutation) - is expected to provide a more holistic model. Permits correlating/inferencing between radiomics and non-radiomics features | + 1 | 1 |
| 7. Detect and discuss biological correlates - demonstration of phenotypic differences (possibly associated with underlying gene–protein expression patterns) deepens understanding of radiomics and biology | + 1 | 1 |
| 8. Cut-off analyses - determine risk groups by either the median, a previously published cut-off or report a continuous risk variable. Reduces the risk of reporting overly optimistic results | + 1 | 1 |
| 9. Discrimination statistics - report discrimination statistics (for example, C-statistic, ROC curve, AUC) and their statistical significance (for example, p-values, confidence intervals). One can also apply resampling method (for example, bootstrapping, cross-validation) | + 1 (if a discrimination statistic and its statistical significance are reported) + 1 (if a resampling method technique is also applied) | 2 |
| 10. Calibration statistics - report calibration statistics (for example, Calibration-in-the-large/slope, calibration plots) and their statistical significance | + 1 (if a calibration statistic and its statistical significance are reported) + 1 (if a resampling method technique is also | 2 |
| (for example, *P*-values, confidence intervals). One can also apply resampling method (for example, bootstrapping, cross-validation) | applied) |  |
| 11. Prospective study registered in a trial database - provides the highest level of evidence supporting the clinical validity and usefulness of the radiomics biomarker | + 7 (for prospective validation of a radiomics signature in an appropriate trial) | 0 |
| 12. Validation - the validation is performed without retraining and without adaptation of the cut-off value, provides crucial information with regard to credible clinical performance | - 5 (if validation is missing) + 2 (if validation is based on a dataset from the same institute) + 3 (if validation is based on a dataset from another institute) + 4 (if validation is based on two datasets from two distinct institutes) + 4 (if the study validates a previously published signature) + 5 (if validation is based on three or more datasets from distinct institutes)<br>*Datasets should be of comparable size and should have at least 10 events per model feature | 4 |
| 13. Comparison to 'gold standard' - assess the extent to which the model agrees with/is superior to the current 'gold standard' method (for example, TNM-staging for survival prediction). This comparison shows the added value of radiomics | + 2 | 2 |
| 14. Potential clinical utility - report on the current and potential application of the model in a clinical setting (for example, decision curve analysis). | + 2 | 2 |
| 15. Cost-effectiveness analysis - report on the cost-effectiveness of the clinical application (for example, QALYs generated) | + 1 | 0 |
| 16. Open science and data - make code and data publicly available. Open science facilitates knowledge transfer and reproducibility of the study | + 1 (if scans are open source) + 1 (if region of interest segmentations are open source) + 1 (if code is open source) + 1 (if radiomics features are calculated on a set of representative ROIs and the calculated features and representative ROIs are open source) | 0 |
| Total points (36 = 100%) | 36 | 18 (50%) |

**Table 2.** TRIPOD recommendations. An “X” indicates if the objective is addressed in the study:

| Section/Topic | n | Checklist Item | Page |
| --- | --- | --- | --- |
| **Title and abstract CT Radiomics Models for Prognostication and Risk-Stratification of Recurrence & Death after Curative-Intent Radiotherapy for Non-Small Cell Lung Cancer** |  |  |  |
| Title | 1 | Identify the study as developing and/or validating a multivariable prediction model, the target population, and the outcome to be predicted. | X |
| Abstract | 2 | Provide a summary of objectives, study design, setting, participants, sample size, predictors, outcome, statistical analysis, results, and conclusions. | X |
| **Introduction** |  |  |  |
| Background and objectives | 3a | Explain the medical context (including whether diagnostic or prognostic) and rationale for developing or validating the multivariable prediction model, including references to existing models. | X |
|  | 3b | Specify the objectives, including whether the study describes the development or validation of the model or both. | X |
| **Method** |  |  |  |
| Source of data | 4a | Describe the study design or source of data (e.g., randomized trial, cohort, or registry data), separately for the development and validation datasets, if applicable. | X |
|  | 4b | Specify the key study dates, including start of accrual; end of accrual; and, if applicable, end of follow-up. | X |
| Participants | 5a | Specify key elements of the study setting (e.g., primary care, secondary care, general population) including number and location of centres. | X |
|  | 5b | Describe eligibility criteria for participants. | X |
|  | 5c | Give details of treatments received, if relevant. |  |
| Outcome | 6a | Clearly define the outcome that is predicted by the prediction model, including how and when assessed. | X |
|  | 6b | Report any actions to blind assessment of the outcome to be predicted. |  |
| Predictors | 7a | Clearly define all predictors used in developing or validating the multivariable prediction model, including how and when they were measured. | X |
|  | 7b | Report any actions to blind assessment of predictors for the outcome and other predictors. |  |
| Sample size | 8 | Explain how the study size was arrived at. | X |
| Missing data | 9 | Describe how missing data were handled (e.g., complete-case analysis, single imputation, multiple imputation) with details of any imputation method. | X |
| Statistical analysis methods | 10a | Describe how predictors were handled in the analyses. | X |
|  | 10b | Specify type of model, all model-building procedures (including any predictor selection), and method for internal validation. | X |
|  | 10d | Specify all measures used to assess model performance and, if relevant, to compare multiple models. | X |
| Risk groups | 11 | Provide details on how risk groups were created, if done. | X |
| **Results** |  |  |  |
| Participants | 13a | Describe the flow of participants through the study, including the number of participants with and without the outcome and, if applicable, a summary of the follow-up time. A diagram may be helpful. | X |
|  | 13b | Describe the characteristics of the participants (basic demographics, clinical features, available predictors), including the number of participants with missing data for predictors and outcome. | X |
| Model development | 14a | Specify the number of participants and outcome events in each analysis. | X |
|  | 14b | If done, report the unadjusted association between each candidate predictor and outcome. | X |
| Model specification | 15a | Present the full prediction model to allow predictions for individuals (i.e., all regression coefficients, and model intercept or baseline survival at a given time point). |  |
|  | 15b | Explain how to use the prediction model. |  |
| Model performance | 16 | Report performance measures (with CIs) for the prediction model. | X |
| **Discussion** |  |  |  |
| Limitations | 18 | Discuss any limitations of the study (such as nonrepresentative sample, few events per predictor, missing data). | X |
| Interpretation | 19b | Give an overall interpretation of the results, considering objectives, limitations, and results from similar studies, and other relevant evidence. | X |
| Implications | 20 | Discuss the potential clinical use of the model and implications for future research. | X |

| Other information |  |  |  |
| --- | --- | --- | --- |
| Supplementary information | 21 | Provide information about the availability of supplementary resources, such as study protocol, Web calculator, and datasets. | X |
| Funding | 22 | Give the source of funding and the role of the funders for the present study. | X |

**Table 3.** univariate Cox Proportional Hazzard Models for *CCNE1*, *BRCA1*, *BRCA2*.

| Data | Metric | Model |  |  |
| --- | --- | --- | --- | --- |
|  |  | *CCNE1* | *BRCA1* | *BRCA2* |
| HH | C-index | 0.52 ± 0.14 | 0.58 ± 0.16 | 0.52 ± 0.13 |
|  | tAUC | 0.50 ± 0.012 | 0.54 ± 0.011 | 0.50 ± 0.012 |
|  | HR | 0.61 (0.1- 3.301) | 3.522<br>(0.02- 4.32) | 1.51 (0.0073-3.17 |
|  | P-value | 0.60 | 0.60 | 0.91 |
| TCIA | C-index | 0.46 ± 0.072 | 0.56 ± 0.064 | 0.55 ± 0.081 |
|  | tAUC | 0.53 ± 0.023 | 0.52 ± 0.012 | 0.47 ± 0.032 |
|  | HR | 0.97 (0.53-1.79) | 1.18 (0.31-4.53) | 0.8 (0.24- 2.70) |
|  | P-value | 0.01 | 0.80 | 0.70 |

**Table 4.** Top 10 most correlated RPPA variables with Spearman’s Rho and P-values (TCIA cohort).

| Protein | Spearman Rho | P-value |
| --- | --- | --- |
| STAT5ALPHA | 0.45 | 0.0016 |
| HER3PY1298 | -0.41 | 0.0040 |
| MTORPS2448 | -0.37 | 0.0098 |
| RAD51.x | -0.37 | 0.011 |
| CHK1PS345 | -0.35 | 0.016 |
| SHCPY317 | -0.31 | 0.032 |
| EGFRPY1068 | -0.31 | 0.034 |
| CDK1.x | -0.30 | 0.037 |
| CIAP | -0.29 | 0.049 |
| AKTPT308 | -0.28 | 0.051 |

**Table 5.**
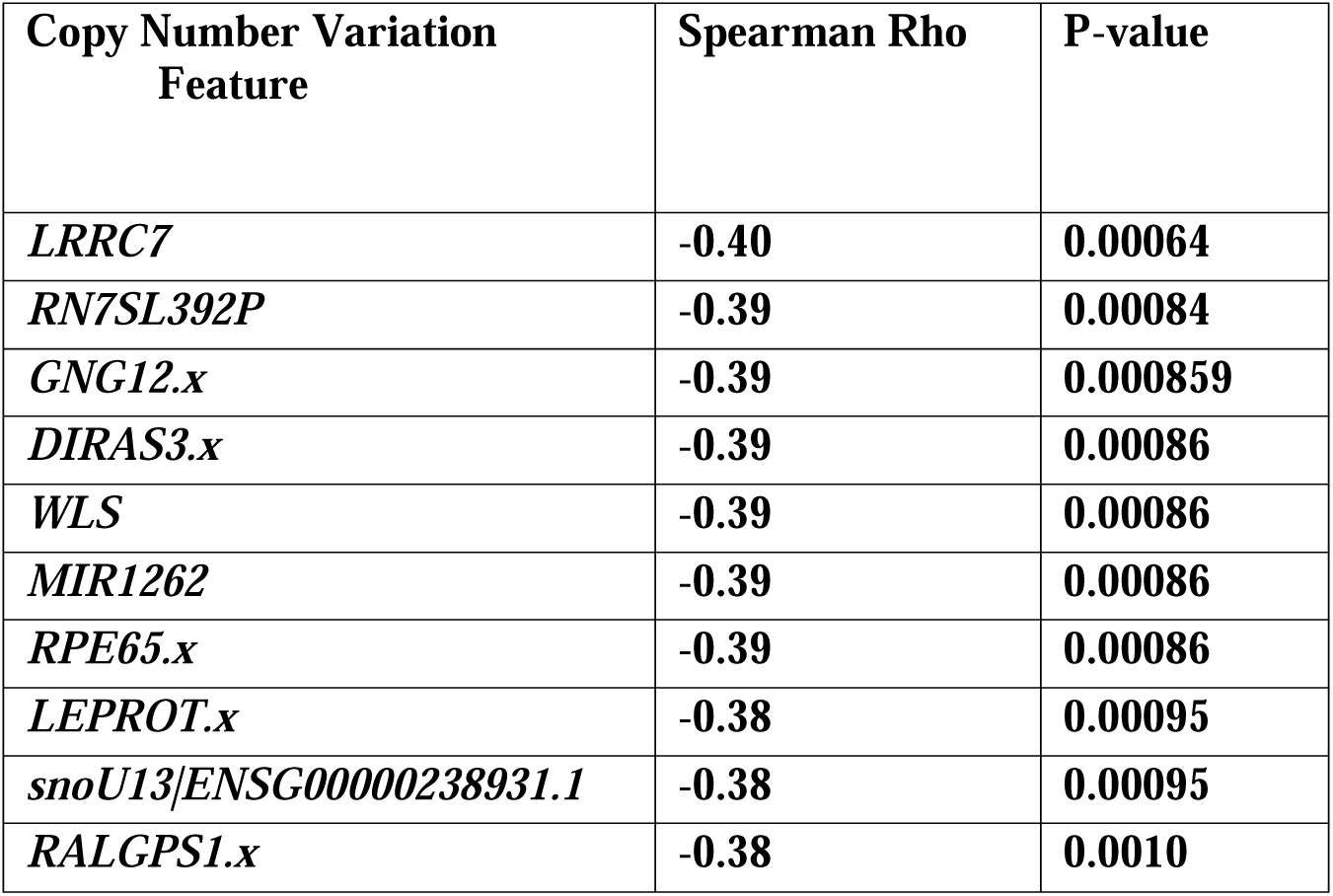
Top 10 most correlated Copy Number Variation variables with Spearman’s Rho and P-values (TCIA cohort)

| Copy Number Variation Feature | Spearman Rho | P-value |
| --- | --- | --- |
| *LRRC7* | -0.40 | 0.00064 |
| *RN7SL392P* | -0.39 | 0.00084 |
| *GNG12.x* | -0.39 | 0.000859 |
| *DIRAS3.x* | -0.39 | 0.00086 |
| *WLS* | -0.39 | 0.00086 |
| *MIR1262* | -0.39 | 0.00086 |
| *RPE65.x* | -0.39 | 0.00086 |
| *LEPROT.x* | -0.38 | 0.00095 |
| *snoU13\|ENSG00000238931.1* | -0.38 | 0.00095 |
| *RALGPS1.x* | -0.38 | 0.0010 |

**Table 6.** Top 10 most correlated gene expression variables with Spearman’s Rho and P-values (TCIA cohort)

| RNA<br>vari<br>able | Spearman Rho | P-value |
| --- | --- | --- |
| *TF.y* | -0.46 | 7.85E-05 |
| *FNDC4.y* | -0.45 | 1.47E-04 |
| *ZNF671.y* | -0.42 | 3.38E-04 |
| *PC.y* | -0.41 | 4.68E-04 |
| *FAAH.y* | -0.40 | 7.74E-04 |
| *MIZF* | -0.40 | 8.08E-04 |
| *SOX30.y* | -0.40 | 8.76E-04 |
| *PTGES2.y* | -0.40 | 8.83E-04 |
| *B3GNT4.y* | -0.39 | 9.52E-04 |
| *FPGS.y* | -0.39 | 1.06E-03 |

**Table 7.** Performance of Radiomics Multi-variable PVIRF-RSF (3 categories variable) and additional clinical variables)

| Data |  | HH | TCIA | KEM |
| --- | --- | --- | --- | --- |
| C-index |  | 0.72 ± 0.068 | 0.74 ± 0.051 | 0.73 ± 0.025 |
| tAUC |  | 0.72 ± 0.04 | 0.74 ± 0.051 | 0.72 ± 0.041 |
| P-value |  | 0.0040 | 0.030 | 2. 01E<sup>-12</sup> |
| HR | Age | 1.001 (0.97 - 1.031) | 1.04 (0.98-1.10) | 1.02 (0.99-2.65) |
|  | Stage | 2.66 (1.30-5.49) | 1.79 (0.81-3.99) | 2.03<br>(1.34-2.07) |
|  | Residual disease | 1.25 (0.53- 2.98) | 1.33<br>( 0.37-4.81) | 3.64 (2.32-5.70) |
|  | PVIRF-RSF | 2.66 (1.30 -5.49) | 3.49 (1.43-8.51) | 1.36 (1.033-1.78) |

